# Lineage replacement and evolution captured by three years of the United Kingdom Covid Infection Survey

**DOI:** 10.1101/2022.01.05.21268323

**Authors:** Katrina Lythgoe, Tanya Golubchik, Matthew Hall, Thomas House, Roberto Cahuantzi, George MacIntyre-Cockett, Helen Fryer, Laura Thomson, Anel Nurtay, Mahan Ghafani, David Buck, Angie Green, Amy Trebes, Paolo Piazza, Lorne J Lonie, Ruth Studley, Emma Rourke, Darren Smith, Matthew Bashton, Andrew Nelson, Matthew Crown, Clare McCann, Gregory R Young, Rui Andre Nunes dos Santos, Zack Richards, Adnan Tariq, Wellcome Sanger Institute COVID-19 Surveillance Team, COVID-19 Infection Survey Group, The COVID-19 Genomics UK (COG-UK) Consortium, Christophe Fraser, Ian Diamond, Jeff Barrett, Ann Sarah Walker, David Bonsall

## Abstract

The Office for National Statistics COVID-19 Infection Survey (ONS-CIS) is the largest surveillance study of SARS-CoV-2 positivity in the community, and collected data on the United Kingdom (UK) epidemic from April 2020 until March 2023 before being paused. Here, we report on the epidemiological and evolutionary dynamics of SARS-CoV-2 determined by analysing the sequenced samples collected by the ONS-CIS during this period. We observed a series of sweeps or partial sweeps, with each sweeping lineage having a distinct growth advantage compared to their predecessors. The sweeps also generated an alternating pattern in which most samples had either S-gene target failure (SGTF) or non-SGTF over time. Evolution was characterised by steadily increasing divergence and diversity within lineages, but with step increases in divergence associated with each sweeping major lineage. This led to a faster overall rate of evolution when measured at the between-lineage level compared to within lineages, and fluctuating levels of diversity. These observations highlight the value of viral sequencing integrated into community surveillance studies to monitor the viral epidemiology and evolution of SARS-CoV-2, and potentially other pathogens, particularly in the current phase of the pandemic with routine RT-PCR testing now ended in the community.

## Introduction

A crucial component of the global response to COVID-19 has been the identification, tracking and characterisation of new SARS-CoV-2 lineages. As well as enabling researchers to identify patterns of spread, variants can be identified that might pose a particular risk. For instance, they may be able to transmit more easily, or evade immune responses. Prominent examples include the variants of concern (VOCs) Alpha, Beta, Gamma, Delta and Omicron (WHO 2023; UKHSA 2023), and individual mutations such as E484K, an immune escape mutation in the Spike protein (Harvey et al. 2021; Carabelli et al. 2023). As of April 2023, around 3 million SARS-CoV-2 sequences had been generated in the United Kingdom (UK) via the COG-UK Genomics Consortium (The COVID-19 Genomics UK (COG-UK) consortium 2020) and the four UK Public Health Agencies, with this substantial surveillance effort generating a snapshot of the leading edge of infection across the UK.

Estimating the prevalence of SARS-CoV-2 lineages and/or mutations can, however, be subject to biases as a consequence of the sampling regime (Franceschi et al. 2021; Kraemer et al. 2019; Pouwels et al. 2021; Mohanan et al. 2021; Wu et al. 2020; Oude Munnink et al. 2021). Sampling has been heavily focussed on symptomatic infections, even though a high proportion of infections are asymptomatic or may not reach the criteria for testing (Sah et al. 2021). For example, in the early phase of the UK epidemic most testing was conducted among hospitalised patients with severe disease, with a later focus on symptomatic individuals. Where testing of asymptomatic individuals has been conducted, it has often been in the context of specific settings, such as returning travellers, schools, or as part of surge testing in geographical areas where VOCs have been identified (UKHSA 2021). Large-scale community surveillance studies, such as the Office for National Statistics Covid Infection Survey (ONS-CIS) (Pouwels et al. 2021), are thus valuable since sampling is not subject to these biases, they consist of a random, potentially more representative sample of the population, and, crucially, identify both symptomatic and asymptomatic infections. Moreover, community-based surveillance studies are not reliant on sequencing samples collected as part of national RT-PCR testing programmes and may therefore become increasingly important as countries seek to enhance surveillance capabilities for SARS-CoV- 2 and other pathogens.

The ONS-CIS is a UK household-based surveillance study, with households approached from address lists to ensure as representative a sample of the population as possible (Pouwels et al. 2021; Office for National Statistics 2021a). Here, we present an analysis of over 125,000 good quality consensus sequences from RT-PCR positive samples collected over the first three years of the survey with the aim of reconstructing the key epidemiological and evolutionary features of the UK epidemic. These data captured the sequential sweeps and partial sweeps of the B.1.177, B.1.1.7/Alpha, B.1.617.2/Delta, and Omicon (BA.1, BA.2, BA.4, BA.5) lineages, the BA.2 recombinant lineage XBB, and BA.2.75 (a sublineage of BA.2) and BQ.1 (a sublineage of BA.5). For each sweeping lineage, we calculated the growth rate advantage using a novel method based on Gaussian processes. This method has the benefit of providing smooth estimates for prevalence and growth rates at both the very low and sometimes zero case counts observed for some variants when they first emerge, and the very high counts once they are established. We also captured the curious alternation of sweeping lineages that exhibit RT-PCR S-gene target failure (SGTF) caused by the Spike DH69/V70 deletion, with those without the deletion, a pattern also observed in other countries such as South Africa (Tegally et al. 2022).

Finally, we determined how these sweeps impacted measures of the genetic diversity and divergence of the virus, both at the within-lineage and between-lineage levels. Using only samples collected as part of the ONS-CIS, we observed a consistent pattern of low within-lineage diversity on first emergence, followed by a steady increase. To capture measures of divergence in a computationally efficient way, we downsampled sequences using weighted random sampling, enabling us to generate a phylogeny in which the major lineages were as evenly distributed through time as possible. Using linear regression we compared the overall rate of divergence with those of the major lineages, with consistently much lower rates of evolution within than between lineages.

Although sequences from the ONS-CIS represented about 4% of the total number of SARS-CoV-2 sequences obtained in the UK during this period, and at times much less than 1%, we were able to reconstruct the key epidemiological and evolutionary aspects of the epidemic. Our observations highlight that representative sampling can capture key aspects of epidemics, and the important role that community-based genomic surveillance studies can have in the monitoring of infectious disease. Although the ONS-CIS is based in the UK, in which sequencing effort has been unprecedented, this is of particular importance in settings where routine testing is likely to be scaled back, and for countries exploring the best strategies for tracking SARS-CoV-2 as well as other respiratory pathogens in the future (Subissi et al. 2022; WHO 2022)

## Results

### Sequential replacement of lineages in the UK

Throughout the ONS-CIS, which was launched in April 2020, a selection of the samples positive by RT-PCR have been sequenced (Figure S1), and from December 2020 onwards the aspiration was to sequence all samples with Ct<=30. Here we report on the sequenced samples with over 50% genome coverage collected between 26th April 2020 and 13th March 2023. The Pango lineage (Rambaut, Holmes, et al. 2020) for all the samples was determined using Pangolin (O’Toole et al. 2021), and from 7 December 2020 onwards we provided publicly available weekly reports, giving the breakdown of sequenced samples (with >50% genome coverage) by lineage (Office for National Statistics 2021b).

The proportion of samples that were RT-PCR positive waxed and waned during the UK epidemic (Figure 1A, S1). There was a small 2020 autumn peak dominated by B.1.177 and its sub-lineages, followed by a decline in cases due to the second national lockdown which lasted from 5th November to 2nd December 2020. Subsequent to this, the number of positives started to rise again. This rise is attributed to a relaxation of restrictions during the Christmas period and corresponded to a rapid rise in the number of B.1.1.7/Alpha infections. After the commencement of a further lockdown in England, Scotland and Northern Ireland in early January 2021, cases declined again, before another rapid increase in the number of sequenced samples that were dominated by B.1.617.2/Delta, with this increase corresponding to a phased reopening on the 19th May and 20th June 2021. Three larger peaks in infections were observed between December 2021 and August 2022, each associated with the sequential emergence of the major Omicron lineages (BA.1, BA.2, and BA.4 and BA.5). Two subsequent smaller waves were not associated with any one lineage, but were dominated by sublineages of BA.2 and BA.5, before a final wave in which XBB, a recombinant BA.2 lineage, gained the advantage. Notably these three final waves all had higher peaks than the earlier B.1.1.77, B.1.1.7/Alpha, and B.1.617.2/Delta waves.

**Figure 1.**
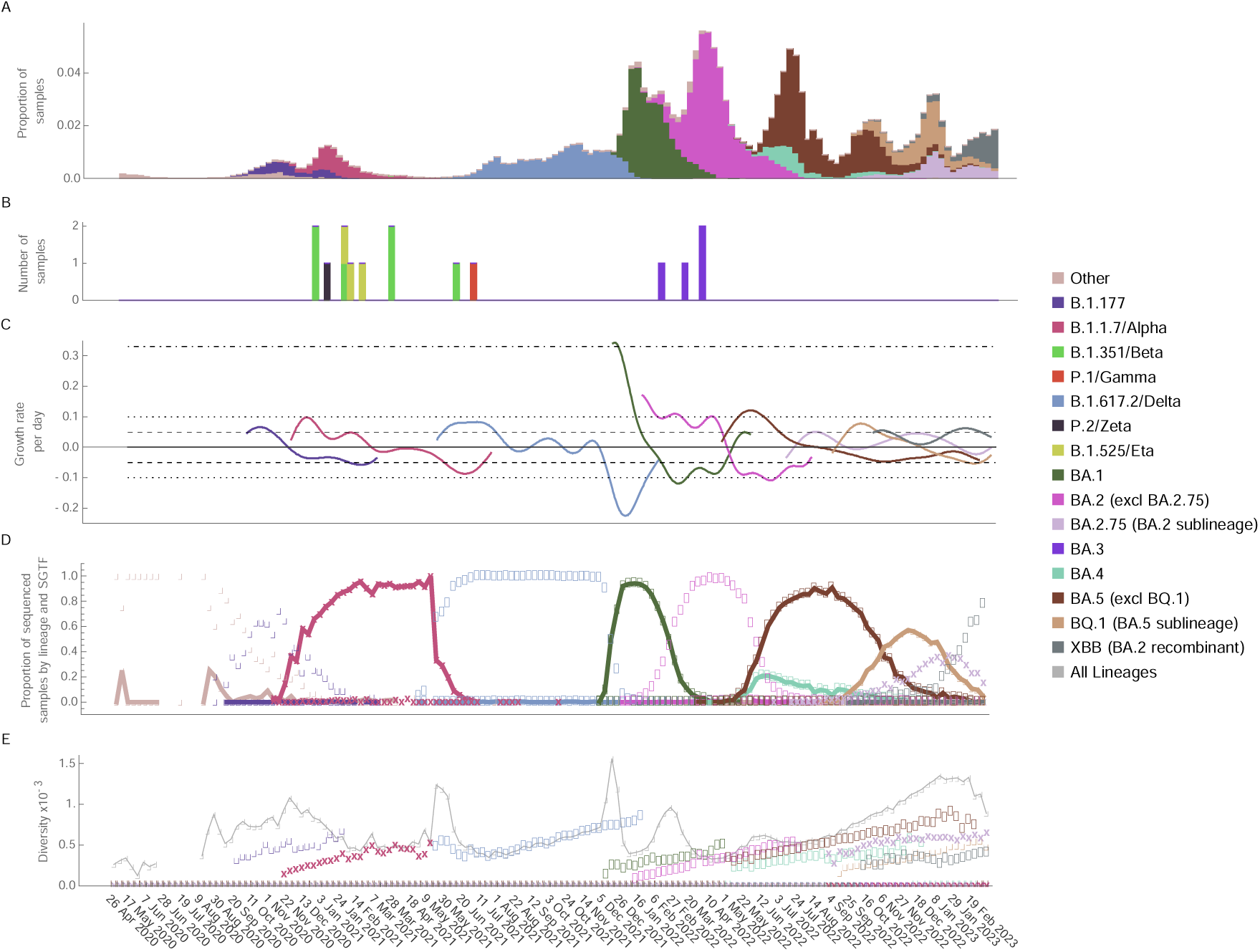
Lineage dynamics and genetic diversity through time. A. Proportion of swabs taken as part of the ONS-CIS that were positive with Ct <=30 (Figure S1), with bars coloured by the proportion of sequenced samples belonging to each major lineage. B. Number of Variant of Concern (VOC) sequenced samples from rarer lineages. C. Per day growth rate advantage of each of the major lineages compared to all other contemporary samples. BA.4 and BA.5 were considered together due to their concordant trajectories, and uncertainty is represented by 200 data bootstraps. The horizontal lines represent how long it would take the VOC prevalence to double (14 days, dashed; 7 days dotted; 2 days dot-dash). D. The bold markers represent the proportion of sequenced samples that were of both the indicated lineage and had S-gene target failure (SGTF) during RT-PCR testing. The pale markers indicate those samples that were non-SGTF. E. Genetic diversity among all samples and among samples of the same major lineages. Lineage designations include all sub-lineages except where indicated, and all samples were grouped by the week in which they were collected, with the date giving the first day of the collection week (every third week labelled for clarity).

### Sweeping lineages each had a substantial growth rate advantage over previously circulating lineages

For each of the sweeping lineages, that is those that rose from low to high frequency, we calculated the relative growth advantage compared to the background of all other lineages that were circulating at the same time (Figure 1C). Because the ONS-CIS data was collected over periods during which the number of positive samples by lineage varied considerably depending on the phase of the epidemic curve, we developed a new method based on Gaussian processes. The code and details for this method are available at https://github.com/thomasallanhouse/covid19-lineages. Importantly, because we compare a specific lineage to all other contemporary lineages, its growth rate advantage will depend on the composition of the background viral population and is not static in time. Hence a lineage which initially enjoys a growth rate advantage will eventually transition to having a disadvantage, as subsequent variants emerge and sweep through the population. Declining growth rate advantages after initial high values have also been noted in previous reports (Kraemer et al. 2021; Volz et al. 2021; Davies, Abbott, et al. 2021; Jones et al. 2021; Vöhringer et al. 2021).

In line with previous findings (Lemey et al. 2021; Hodcroft et al. 2021; Vöhringer et al. 2021; Sonabend et al. 2021; Tegally et al. 2022; Viana et al. 2021) we found that all of the sweeping lineages, including B.1.177, had a significant growth rate advantage compared to all other co-circulating SARS-CoV-2 lineages during emergence, with the maximum per day advantage observed for each major lineage ranging from around 5% (a 14 day doubling time) for B.1.177, BA.2.75/Omicron, and XBB, to around 10% (7 day doubling time) for B.1.1.7/Alpha, B.1.617.2/Delta, BA.2/Omicron, BA.4/5/Omicron, and BQ.1, and around 33% (2 day doubling time) for BA.1/Omicron (Figure 1C).

### Alternating SGTF and non-SGTF

Curiously, the successive sweeps of distinct lineages observed in the UK, as also found in South Africa (Tegally et al. 2022; Viana et al. 2021), have been characterised by the alternation of lineages that exhibit RT-PCR S-gene target failure (SGTF) caused by the Spike DH69/V70 deletion, with those that do not have SGTF (Figure 1D, Figures S1). For all sequenced samples with an assigned lineage, we classed those as having SGTF if, during RT-PCR testing, N and ORF1ab were successfully amplified but S was not, and non-SGTF samples as those where all three genes were amplified. Lineage was generally a good indicator of SGTF, with 98.84% (3416/3456) of B.1.1.7/Alpha, 99.82% (24511/24556) of BA.1/Omicron, 99.77% (3428/3464) of BA.4/Omicron, and 99.53% (27106/27234) of BA.5/Omicron, including BQ.1, samples having SGTF; all four of these lineages have the DH69/V70 deletion. Other lineages, which generally lack the deletion, typically did not have SGTF; 0.48% (7/1450) of B.1.177 samples had SGTF, 0.16% (29/17766) of B.1.617.2/Delta, and 0.53% (226/42938) of BA.2/Omicron, including BA.2.75, and XBB. As previously reported (Thomson et al. 2021), a clear exception was the B.1.258 lineage, of which around two-thirds of samples had both the DH69/V70 deletion and SGTF (48/71), and some B.1.617.2/Delta and BA.2 sequences also had the DH69/V70 deletion, and hence had SGTF (Figure S1).

The independent emergence of DH69/70 on different lineages, including B.1.258, B.1.1.7/Alpha, and BA.1/Omicron, is a prominent example of the convergence that has been observed repeatedly during the evolution of SARS-CoV-2 (McCarthy et al. 2021). This pattern of alternating SGTF greatly facilitated the rapid quantification of the prevalence of different VOCs without the need for genome sequencing, enabling samples with high Ct to be included in epidemiological analyses, and avoiding delays associated with sequencing (Kidd et al. 2021; Walker et al. 2021; Volz et al. 2021; Davies, Jarvis, et al. 2021; Vogels et al. 2021; Tegally et al. 2022; Office for National Statistics 2021c); Intriguingly, the pattern of alternating SGTF and non-SGTF has continued until at least mid-March 2023, even though no new major lineages emerged, with BA.2 recombinant and sublineages that do not have SGTF (including XBB and BA.2.75) gradually outcompeting BA.5 and its sublineages (including BQ.1) which all have SGTF. As of mid-March 2023, XBB was the dominant lineage circulating in the UK.

However, SGTF or non-SGTF has not always been a reliable indicator of the major lineage of a sample. This can be due to i) co-circulation of lineages with the same SGTF pattern such as B.1.358 and B.1.1.7/Alpha; BA.4 and BA.5; and BA.2.75 and XBB; ii) lineages with sub-lineages that have different SGTF patterns, such as B.1.258; and iii) a small proportion of samples being assigned the opposite to expected SGTF pattern - this will have a disproportionate impact when the prevalence of the lineage of interest is low whilst the prevalence of another lineage with the opposite SGTF pattern is high. This highlights the need for caution when using SGTF as a marker for lineage, particularly when prevalences are low, and the necessity of sequencing in obtaining a full understanding of the genetic landscape at any particular time.

### Diversity increases within lineages through time, but fluctuates when measured across all lineages

With the exception of B.1.617.2/Delta, within-lineage diversity was generally low when each major lineage first appeared in the ONS-CIS, and gradually increased through time before replacement by a new variant (Figure 1E). This initial low diversity is a consequence of their relatively recent emergence before first detection in the ONS-CIS data. On the other hand, B.1.617.2/Delta had high initial diversity (Figure 1E), reflecting its repeated introduction from an already diverse source population in India (Public Health England 2021).

In contrast, when we consider overall genetic diversity, we see transient increases, peaking when two or more major lineages are at relatively high frequencies (Figure 1E), but then declining as single lineages dominate the population. This is unsurprising given the large number of mutations distinguishing the different major lineages, which will push up diversity when two or more lineages are relatively frequent. Nonetheless, it is striking that from the Autumn of 2020 to the Spring of 2023 we did not see a trend of increasing levels of global diversity despite nearly two and a half years of evolution during this time. The most sustained period of gradually increasing diversity was during the second half of 2022 as multiple sublineages of BA.2 and BA.5 co-circulated over a prolongued period of time, but this was then followed by a decline as the BA.2 recombinant lineage XBB began to dominate.

### Divergence increases slower within than among lineages

Phylogenies can be difficult to generate for large alignments, although a fast approximate maximum likelihood method has been developed specifically for SARS-CoV-2 (Turakhia et al. 2021). These large phylogenies can also be subject to biases if lineages or epochs are unevenly sampled, and difficult to visualise. To overcome some of these issues, we subsampled 3000 high-coverage (>95%) consensus sequences from the ONS-CIS data using weighted random sampling, so that the VOCs, and the sweeping and partially sweeping lineages, were as similarly represented and as evenly distributed through time as possible. The sequential sweeps of the major lineages are readily observable on the resultant time-scaled phylogeny, with each of the major lineages representing a distinct clade or sub-clade (Figure 2, S2). The sampling methodology meant that lineages that were rarely sampled in the UK, such as B.1.351/Beta, B.1.525/Eta, P.1/Gamma, and BA.3/Omicron, were represented in the phylogeny. Apart from B.1.617.2/Delta, the major UK lineages have times of most recent common ancestor (tMRCAs) close to the time of first sampling in the ONS-CIS, indicating the recent emergence of these lineages when first sampled.

**Figure 2.**
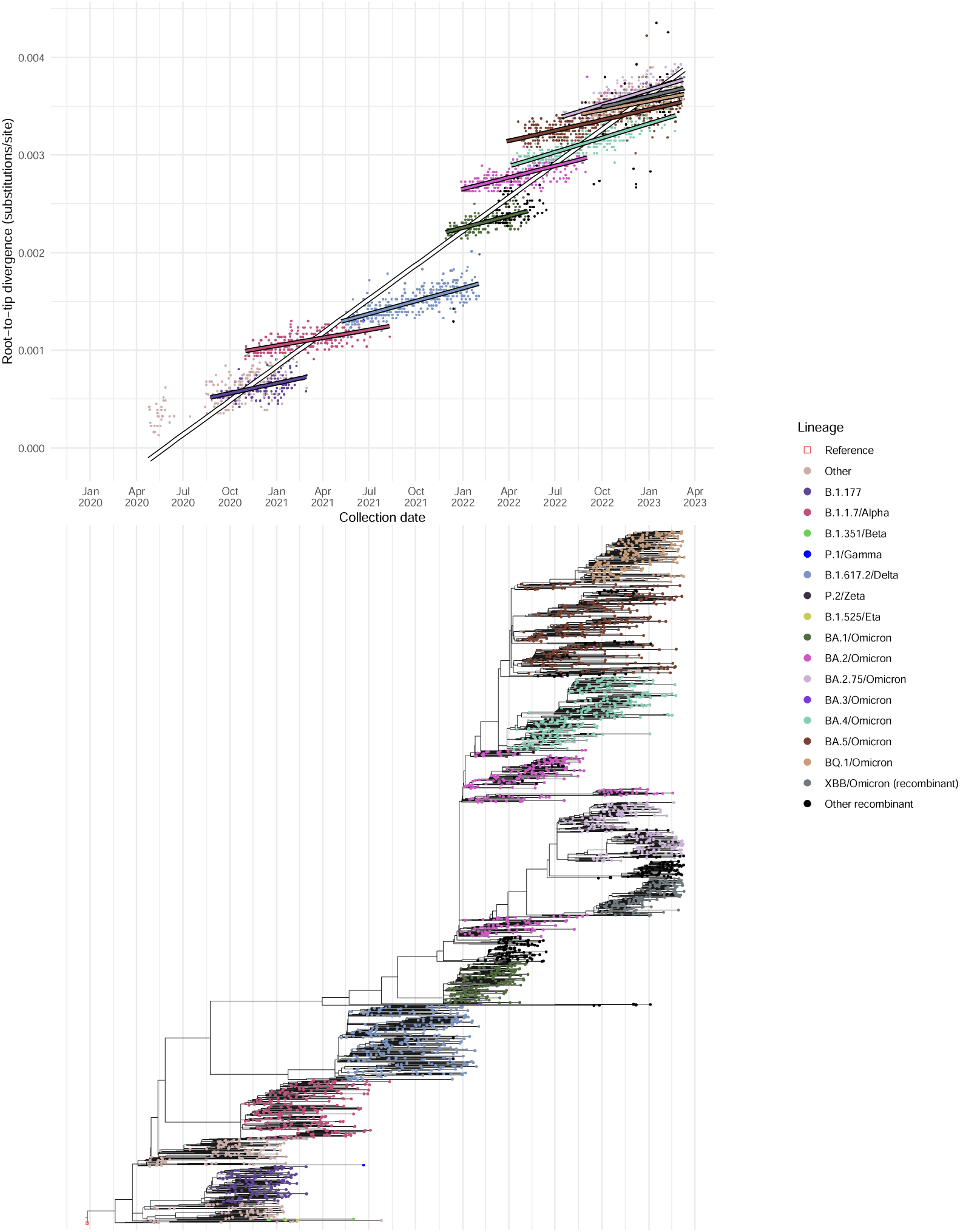
Dated phylogeny and root-to-tip distance of ONS-CIS sequences. A maximum likelihood phylogeny of 3000 ONS sequences with over 95% genome coverage was generated using IQ-TREE (Figure S2). The samples were chosen using a weighted random sampling, ensuring VOCs and other major lineages were as evenly distributed through time as possible Top. Root to tip distance for samples from the maximum likelihood phylogeny. BA.2 sequences (excluding BA.2.75) collected before and after 12th September 2022 were considered separately; the later sequences are all from the BA.2.3.20 lineage or its descendents. Bottom. Time tree generated from the maximum likelihood phylogeny using TreeTime (Sagulenko, Puller, and Neher 2018).

As expected, divergence from the root of the phylogeny increased gradually through time, both within-lineages and across all lineages (Figure 2), demonstrating the presence of a strong molecular clock. The estimated overall mutation rate was 1.39×^-3^ substitutions per site per year by simple linear regression. It has been noted previously that although divergence within the B.1.1.7/Alpha lineage increased at a similar rate to previously circulating lineages, it had accumulated a disproportionate number of lineage defining mutations at the time of emergence (Hill et al. 2022), and that this pattern is also observed for other VOC lineages (Tay et al. 2022; Neher 2022). We can readily see this pattern using the subsampled ONS-sequences, and we formally investigate it using a linear regression of root-to-tip divergence versus calendar time, with lineage used as an interaction term (Table S1). B.1.177, used as the reference category, evolved at an estimated 4.06×^-4^ substitutions per site per year, and there was no evidence that the rate of evolution differed for most other lineages (exceptions being BA.2.75 and BA.4, which were slightly faster).

Substitution rates in the “Other” category, which represents a diverse collection of early sequences and minor lineages, and whose estimate could be seen as representing between-lineage rate, was much faster (1.29×^-3^ subs/site/year). For this analysis, we split the BA.2 samples (excluding BA.2.75) into those collected prior to 12th September 2022 and those after. The later sampled lineages were contemporary to the BA.2.75 wave, and were members of the BA.2 sublineage BA.2.3.20, which harbours a large number of mutations compared to its predecessors, and are observed at a time when all other BA.2 lineages had effectively gone extinct.

The almost one-to-one concordance that we see between the emergence of lineages divergent compared to their predecessors, their growth rate advantage, and the resulting epidemic waves provides a clear narrative of epidemiological dynamics driven by saltational evolutionary events. Since new VOCs and other major lineages are characterised by nonsynonymous mutations (Neher 2022), it has been hypothesised that they arose during long-term chronic infections with the virus subject to strong immune selection (Rambaut, Loman, et al. 2020; Ghafari et al. 2023; Markov et al. 2023).

## Discussion

Surveillance studies are valuable tools for tracking the emergence and spread of an infectious disease. Using sequenced samples collected as part of the ONS-CIS, we have demonstrated the utility of large-scale surveillance studies to identify key epidemiological and evolutionary features of the UK epidemic. Since participating households are chosen to be representative of the UK population, and enrolled individuals are periodically tested for SARS-CoV-2 infection regardless of symptoms, the ONS-CIS gives a picture of SARS-CoV- 2 prevalence in the UK that is not subject to the biases arising from focussing on symptomatic individuals or other groups (Franceschi et al. 2021; Kraemer et al. 2019; Pouwels et al. 2021).

The sequential sweeps of different major lineages in the UK observed during the first three years of the UK epidemic resulted in a pattern of relatively steady within-lineage evolution and gradual increases in within-lineage diversity, followed by step-increases in the number of substitutions as each new major lineage emerged. This in turn produced faster estimates of overall rates of evolution when measured across all lineages, and fluctuating levels of genetic diversity. Whether this pattern will be an ongoing feature of SARS-CoV-2 evolution remains to be seen. However, it is noticeable that even though more recent major lineages have been descendants of previously circulating lineages (BA.2.75 and BA.2.3.20 are both descendents of BA.2, and BQ.1 is descended from BA.5), or recombinants, the large number of mutations these lineages have acquired compared to their predecessors is evident. If the hypothesis that these major lineages arose or partially arose from chronically-infected individuals is correct (Hill et al. 2022; Ghafari et al. 2023), it will be difficult to predict what the next sweeping lineage will look like, as it could potentially be a descendant of Omicron or of one of the earlier circulating lineages, including the more pathogenic B.1.617.2/Delta. This again emphasises the need for effective genomic surveillance at a global scale.

A key observation from our study is that the growth rate advantage for a sweeping lineage will not be static in time. Instead, the relative growth rate of a variant will be determined by a number of factors, including its intrinsic transmissibility and the other viral lineages circulating at the time, but also immune escape and levels of population immunity to all extant variants (Markov et al. 2023). As a result, growth rate advantages may differ by both regions and calendar time, and there should be no *a priori* expectation that variants that have previously disappeared due to competition could not re-emerge, potentially seeded from chronically infected individuals, once the immunological background has changed.

It is tempting to argue that the alternating high prevalences of non-SGTF (B.1.177, B.617.2/Delta, BA.2) and SGTF lineages (B.1.1.7/Alpha, BA.1, BA.4/5), caused by the absence and presence of the DH69/V70 deletion, is a consequence of the changes in the immunological background. DH60/V70 causes conformational changes in the NTD loop and this in turn may compensate for, or have other epistatic interactions with, immune escape mutations (Meng et al. 2021). The gradual increase in the proportion of BA.2 sequences that had DH60/V70 when BA.2 as a whole was in decline, and when BA.4 and BA.5 were beginning to sweep through the population, supports this idea, as does the resurgence of non-SGTF BA.2 recombinant and sub-lineages in the UK population towards the end of 2022, coinciding with a fall in BA.5 lineages. However, without a clear mechanism causing the switching, this remains a hypothesis only, and it is hard to predict whether the pattern will continue into the future.

A crucial drawback to genomic surveillance is the delay between sample collection and subsequent sequencing (in the ONS-CIS this was between two and three weeks) and the need for high viral load samples to produce adequate sequence data. This in turn could impact the success of any interventions. The earliest signals that both B.1.1.7/Alpha and BA.1/Omicron (Viana et al. 2021) had a growth rate advantage were serendipitously inferred from the increasing incidence of SGTF during RT-PCR testing, and conversely increasing non-SGTF was observed when B.1.617/Delta and BA.2 were emerging. However, the simultaneous rise in BA.4 and BA.5, both with SGTF, required sequencing to distinguish between them, and similarly the BA.2 recombinant and sublineages that subsequently overtook BA.4 and BA.5 cannot be distinguished in this way. The introduction of qPCR-based genotyping for specific lineages into diagnostic pipelines has the potential to speed-up detection of known variants (Vogels et al. 2021), but the lead time required to manufacture them means they cannot be relied upon to characterise emerging variants fast enough to contain them. Moreover, substantial genome sequencing efforts will always be required to detect variants that have not previously been identified as of concern, to monitor the ongoing specificity of rapid genotyping in the face of ongoing evolution, and to better characterise the evolution and spread of the virus.

Although only a fraction of the COG-UK sequences were comprised of samples collected as part of the ONS-CIS, we were able to use ONS-CIS sequenced samples to monitor the emergence, spread and evolution of the major lineages and sublineages sweeping through the UK population. Moving forwards, the implementation of genomic surveillance globally should be considered a key development goal, enabling the early detection of worrisome and/or rapidly growing lineages wherever they emerge. Whilst community surveillence at the scale of the ONS-CIS is unlikely to be feasible in most regions of the world, the survey is an important benchmark, and provides data and lessons to be learnt for when designing surveillance studies in other countries. Incorporating the detection and sequencing of other pathogens into the same community surveillance frameworks will only act to enhance the positive public health and scientific outcomes from these studies whilst maximising value for money.

## Methods

### ONS COVID-19 Infection Survey

The ONS-CIS is a UK household-based surveillance study whose participant households are chosen to provide a representative sample of the population. For a full description of the sampling design see (Pouwels et al. 2021), but in brief, swabs were taken from individuals aged two years and older, living in private households, from 26th April 2020 to the 13th March 2023. These households were selected randomly from address lists and previous ONS surveys to provide a representative sample of the population. Participants could provide consent for optional follow-up sampling weekly for the first five weeks, and monthly thereafter.

This work contains statistical data from ONS which is Crown Copyright. The use of the ONS statistical data in this work does not imply the endorsement of the ONS in relation to the interpretation or analysis of the statistical data. This work uses research datasets which may not exactly reproduce National Statistics aggregates.

### Sequencing

A selection of RT-PCR samples were sequenced each week, with some additional retrospective sequencing of stored samples. From December 2020 onwards, the ambition was to sequence all positive samples with Ct<=30. Most samples were sequenced on Illumina Novaseq, but with a small number using Oxford Nanopore GridION or MINION. One of two protocols were used: either the ARTIC amplicon protocol (COG-UK 2020) with consensus FASTA sequence files generated using the ARTIC Nextflow processing pipeline (COG-UK 2020), or veSeq, an RNASeq protocol based on a quantitative targeted enrichment strategy (Lythgoe et al. 2021; Bonsall et al. 2020) with consensus sequences produced using *shiver* (Wymant et al. 2018). For veSeq we have previously shown that viral load is positively correlated with the number of mapped reads, and that Ct is negatively correlated with the Log10 number of mapped reads (Lythgoe et al. 2021; Golubchik et al. 2021; Fryer et al. 2022). Where there was duplicate sequencing, either of the same sample or of multiple samples taken from an individual at the same time, only the sequence with the highest coverage was kept. The proportion of swabs RT-PCR positive, RT-PCR positive and with Ct<=30, and RT-PCR positive, Ct<=30 and sequenced with >50% genome coverage is shown in Figure S1.

### Lineage calling

Lineages using the Pango nomenclature (Rambaut, Holmes, et al. 2020) were determined using the Pangolin software (O’Toole et al. 2021). Reported major lineages include any sub- lineages, with the exception of BA.2, which did not include BA.2.75 and sublineages of BA.2.75, and BA.5 which did not BQ.1 and sublineages of BQ.1. When comparing SGTF with lineage, we excluded samples where the lineage resolved to A, B, B.1 or B.1.1 since sequences can be given these Pango lineages if an insufficient number of loci have coverage at lineage defining sites. The COG-UK ID, collection dates, Pango lineage, major lineage for all sequences included in this study can be downloaded from https://github.com/katrinalythgoe/ONSLineages. All sequences have been made publicly available as part of the COG-UK consortium. The sequences have also been deposited in the European Nucleotide Archive (ENA) at EMBL-EBI as part of the COG-UK consortium, which has accession number PRJEB37886 (https://www.ebi.ac.uk/ena/browser/view/PRJEB37886).

### Lineage growth rates and doubling times

For each of the major lineages, sublineages and recombinant lineages observed at high frequency, B.1.177, B.1.1.7/Alpha, B.1.617.2/Delta, the Omicron variants BA.1, BA.2, and BA.4 and BA.5, BA.2.75, BQ.1 and XBB, we calculated their relative growth rate advantage compared to all other lineages. BA.4 and BA.5 were considered together because of their concordant dynamics. We used a combination of Gaussian process regression and classification (Rasmussen and Williams 2005) together with bootstrapping as detailed fully at https://github.com/thomasallanhouse/covid19-lineages.

### Nucleotide genetic diversity

Nucleotide genetic diversity was calculated using the *π* statistic, since this has been shown to be the least sensitive to differences in the number of sequences used in the analysis (Zhao and Illingworth 2019). Mean pairwise genetic diversity across the genome is given by:

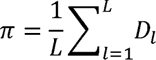

Where *L* represents the length of the genome, and *D_i_* the pairwise genetic diversity at locus *l*. This is calculated as:

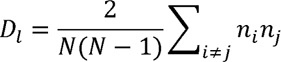

Where *n_i_* represents the number of alleles *i* observed at that locus, and *N* the number of samples with a consensus base call. Within-lineage genetic diversity was similarly calculated as above, but limiting only to the major lineages.

### Phylogenetics

For the phylogenetic analysis, 3000 consensus sequences with at least 95% coverage were chosen using weighted random sampling, with each sample of major lineage *i* collected in week *j* given a weight *1/x_ij_*, where *x_ij_* is the number of sequences of major lineage *i* collected during week *j*. The major lineages included B.1.177, all of the identified VOCs (with BA.1 - BA.5 each given their own weighting), BA.2.75, BQ.1 and XBB. All other well-resolved lineages were placed in the category ‘Other’. The sub-sampled sequences are indicated in the table deposited at https://github.com/katrinalythgoe/ONSLineages. All were pairwise aligned to the Wuhan-Hu-1 reference strain using MAFFT (Katoh and Standley 2013) and then combined to generate a single alignment of 29903 base pairs.

The alignment of 3000 sequences, combined with Wuhan-Hu-1, were used for phylogenetic reconstruction using IQ-TREE version 1.6.12 (Kozlov et al. 2019). The substitution model used was GTR+F+R4 with four FreeRate rate categories, and the resulting tree was rooted using Wuhan-1 and then fit to the calendar using TreeTime version 0.8.2 (Sagulenko, Puller, and Neher 2018). Tips representing the reference strain and sequences judged by TreeTime as outliers on the root-to-tip divergence plot were pruned from the phylogeny and excluded from further analysis. Visualisation used ggtree (Yu, Smith, and Tsan-Yuk 2017).

A linear regression analysis of root-to-tip divergence as a function of time since 1 January 2020 (in decimal years) and lineage was conducted. B.1.177 was used as the baseline category for lineage (Table S1). This included an interaction term for the two dependent variables. For this analysis, the BA.2 lineage was split into two (one for sequences sampled before the 12th September 2022 and one for those after) and any lineages with less than ten examples amongst the 3000 were combined with the “Other” category.

## Data Availability

A privacy preserving version of the lineage dataset is available from ons.gov.uk. The sequences are publicly available via the COG-UK project on the ENA. A table (SampleInfo.csv) has been uploaded to Katrina Lythgoe's Github with the COG-UK ids, enabling them to be identified and downloaded. The table also gives their collection dates, pangolin designation, major lineage, and whether they were included in the phylogenetic analysis. The weekly aggregated ONS-CIS testing data (TestingInfo.csv) is also available here. The full description and code for the growth rate analysis is on Thomas House's github. We have permission from the ONS to make this data publicly available.

https://www.ons.gov.uk/peoplepopulationandcommunity/healthandsocialcare/conditionsanddiseases/datasets/covid19infectionsurveytechnicaldata/2021

https://www.ebi.ac.uk/ena/browser/view/PRJEB37886

https://github.com/katrinalythgoe/ONSLineages

https://github.com/thomasallanhouse/covid19-lineages

---

**Figure S1.**
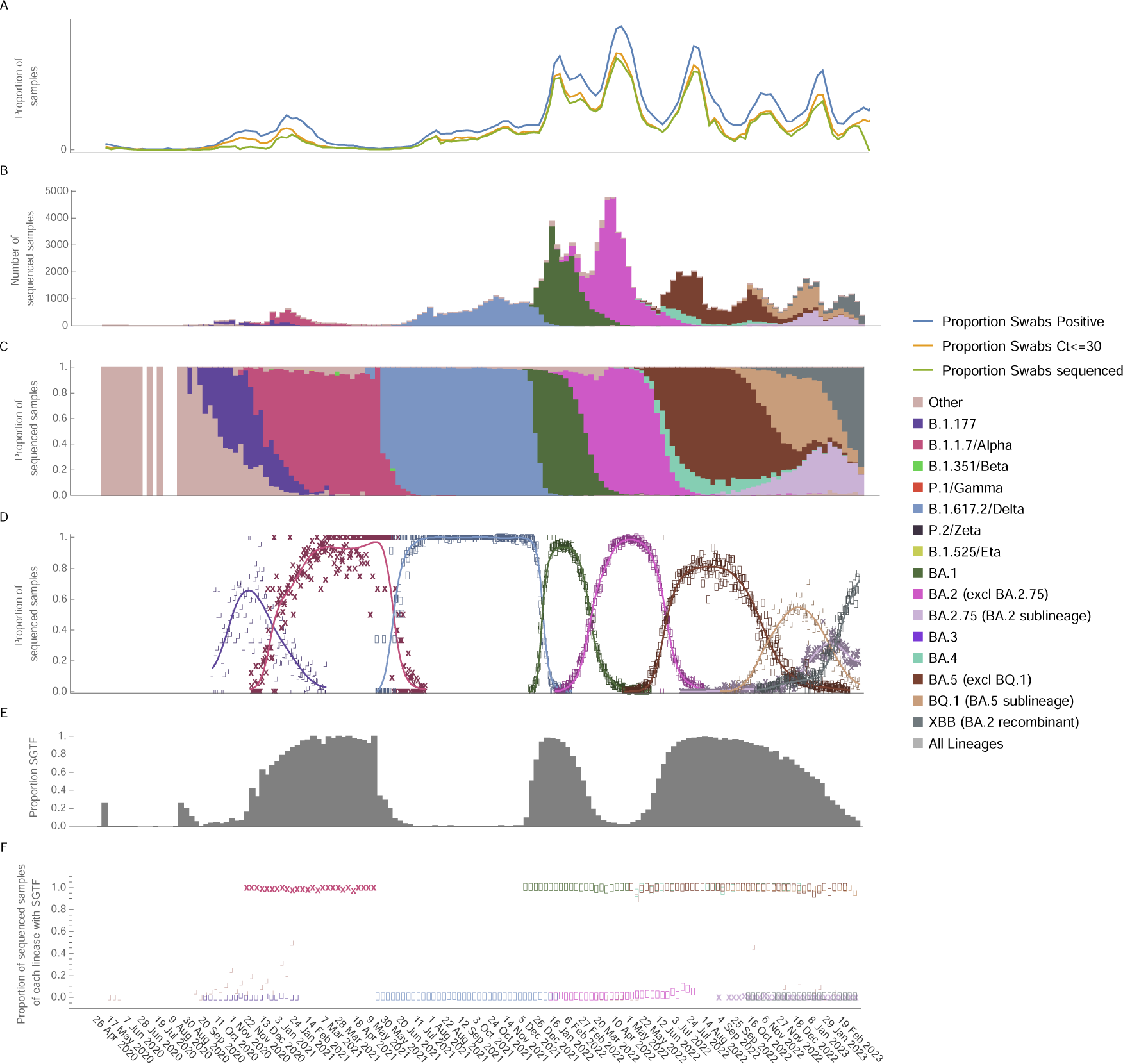
Number and proportion of samples sequenced, lineage dynamics, and patterns of S-gene target failure (SGTF). A. Proportion of swabs RT-PCR positive; RT-PCR positive and with Ct<=30; RT-PCR positive, Ct<=30, and sequenced with >50% genome coverage. B. Number of sequenced samples from the ONS-CIS coloured by major lineage and grouped by collection week. C. Proportion of sequenced samples of each major lineage with samples grouped by collection week. D. The proportion of samples belonging to each of the lineages compared to all other contemporary samples on each day of sampling, with uncertainty represented by 200 data bootstraps. For this analysis, BA.4 and BA.5 were considered together (dark brown) due to their concordant dynamics. E. Proportion of sequenced samples with RT-PCR SGTF. F. Proportion of samples, for the each of the major lineages, that had SGTF. Every third week is labelled for clarity.

**Figure S2.**
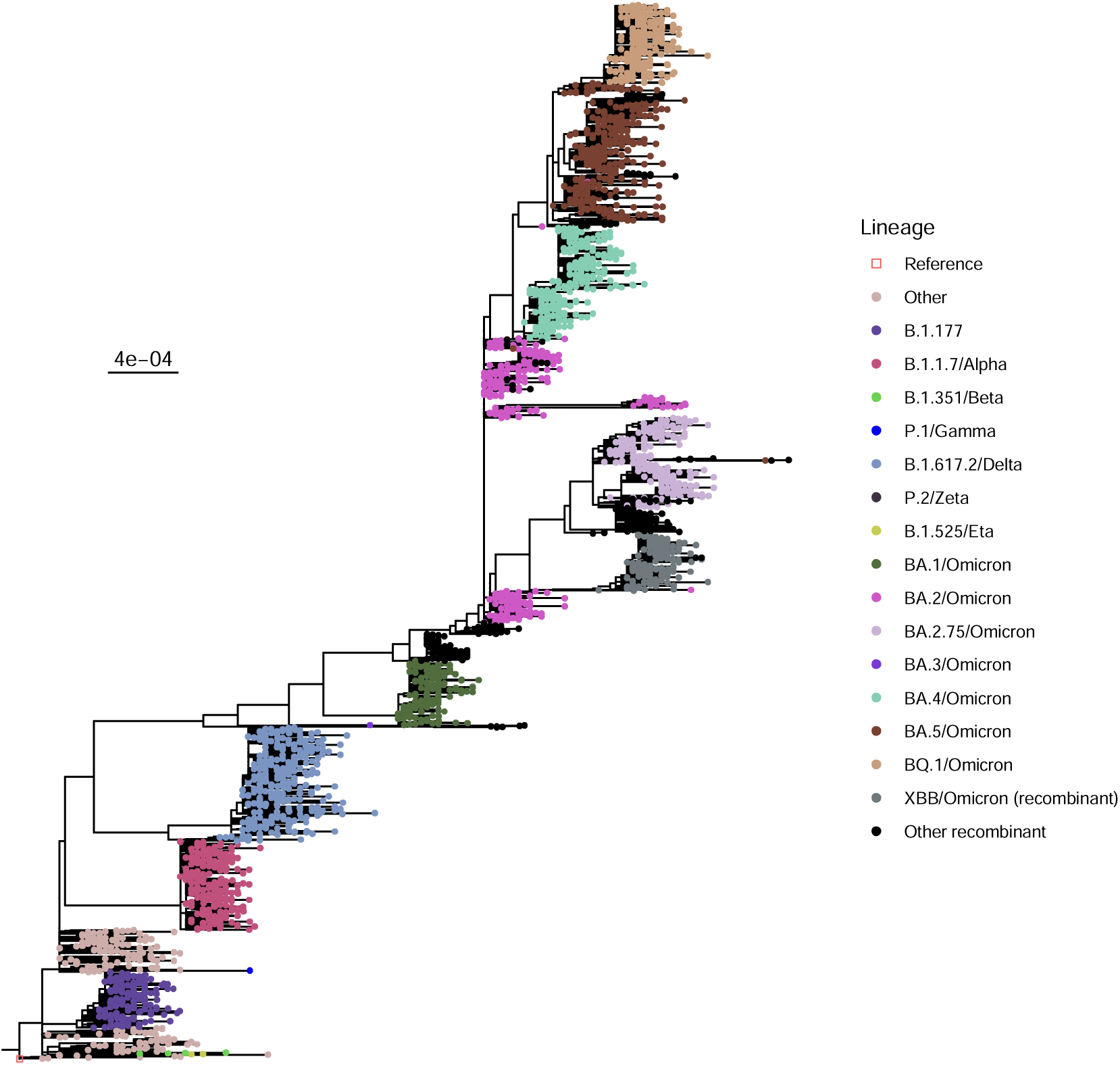
Maximum likelihood phylogeny of ONS-CIS sequences. A maximum likelihood phylogeny of 3000 ONS sequences with over 95% genome coverage was generated using IQ-TREE. The samples were chosen using weighted random sampling, ensuring VOCs and other major lineages were as evenly distributed through time as possible.

**Table S1:**
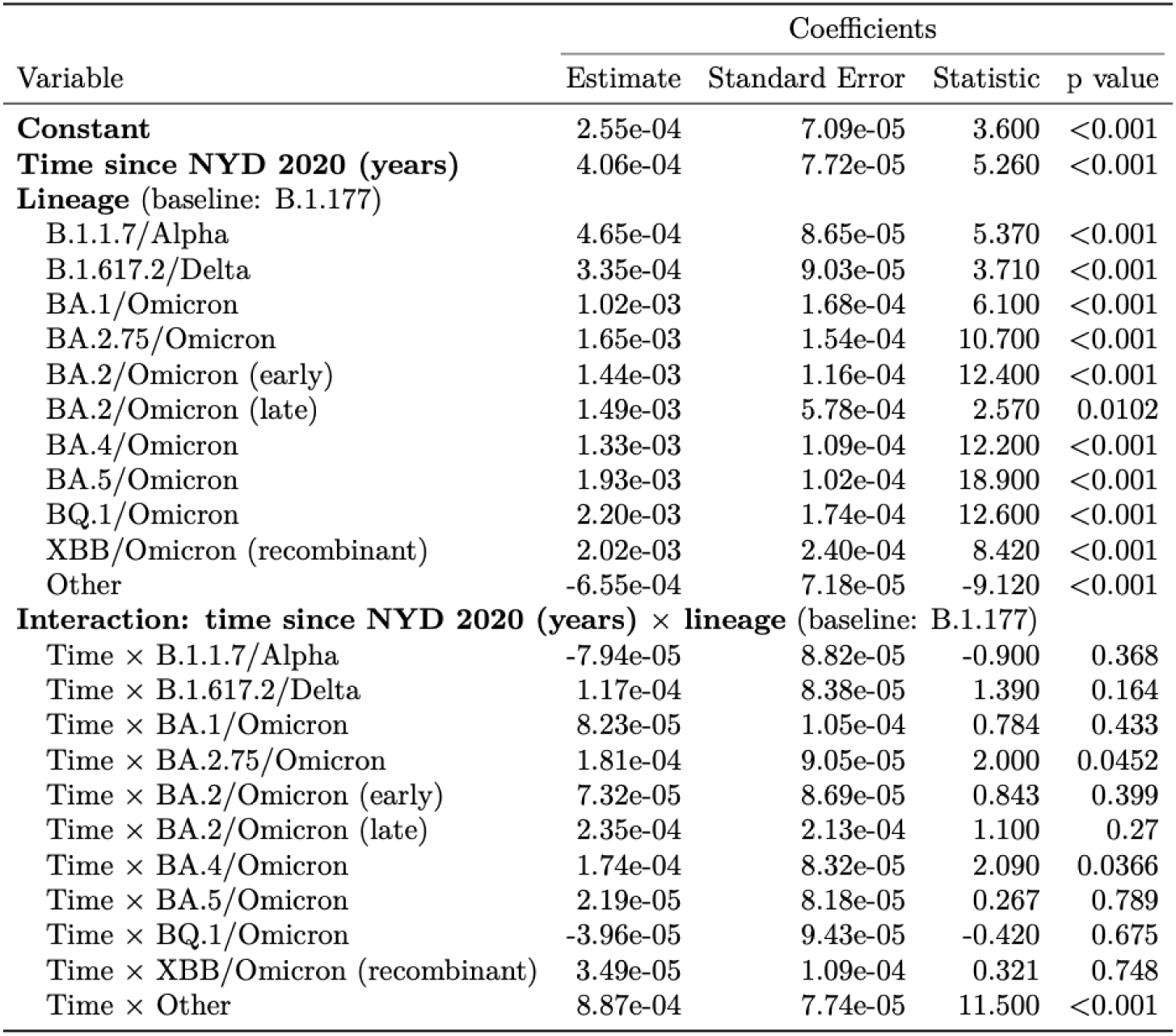
Linear regression of root-to-tip divergence as a function of time since 1 January 2020 (in decimal years) and lineage Time since NYD (New Year’s Day, 1 January) 2020 gives the substitution rate for the B.1.177 lineage (substitutions per site per year), and the intercept gives where the regression line crosses the y-intercept. The lineage estimate gives the addition to the B.1.177 intercept for subsequence lineages, and the interaction estimate gives the addition to the substitution rate.

| Variable | Coefficients |  |  |  |
| --- | --- | --- | --- | --- |
|  | Estimate | Standard Error | Statistic | p value |
| **Constant** | 2.55e-04 | 7.09e-05 | 3.600 | <0.001 |
| **Time since NYD 2020 (years)** | 4.06e-04 | 7.72e-05 | 5.260 | <0.001 |
| **Lineage** (baseline: B.1.177) |  |  |  |  |
| B.1.1.7/Alpha | 4.65e-04 | 8.65e-05 | 5.370 | <0.001 |
| B.1.617.2/Delta | 3.35e-04 | 9.03e-05 | 3.710 | <0.001 |
| BA.1/Omicron | 1.02e-03 | 1.68e-04 | 6.100 | <0.001 |
| BA.2.75/Omicron | 1.65e-03 | 1.54e-04 | 10.700 | <0.001 |
| BA.2/Omicron (early) | 1.44e-03 | 1.16e-04 | 12.400 | <0.001 |
| BA.2/Omicron (late) | 1.49e-03 | 5.78e-04 | 2.570 | 0.0102 |
| BA.4/Omicron | 1.33e-03 | 1.09e-04 | 12.200 | <0.001 |
| BA.5/Omicron | 1.93e-03 | 1.02e-04 | 18.900 | <0.001 |
| BQ.1/Omicron | 2.20e-03 | 1.74e-04 | 12.600 | <0.001 |
| XBB/Omicron (recombinant) | 2.02e-03 | 2.40e-04 | 8.420 | <0.001 |
| Other | -6.55e-04 | 7.18e-05 | -9.120 | <0.001 |
| **Interaction: time since NYD 2020 (years) × lineage** (baseline: B.1.177) |  |  |  |  |
| Time × B.1.1.7/Alpha | -7.94e-05 | 8.82e-05 | -0.900 | 0.368 |
| Time × B.1.617.2/Delta | 1.17e-04 | 8.38e-05 | 1.390 | 0.164 |
| Time × BA.1/Omicron | 8.23e-05 | 1.05e-04 | 0.784 | 0.433 |
| Time × BA.2.75/Omicron | 1.81e-04 | 9.05e-05 | 2.000 | 0.0452 |
| Time × BA.2/Omicron (early) | 7.32e-05 | 8.69e-05 | 0.843 | 0.399 |
| Time × BA.2/Omicron (late) | 2.35e-04 | 2.13e-04 | 1.100 | 0.27 |
| Time × BA.4/Omicron | 1.74e-04 | 8.32e-05 | 2.090 | 0.0366 |
| Time × BA.5/Omicron | 2.19e-05 | 8.18e-05 | 0.267 | 0.789 |
| Time × BQ.1/Omicron | -3.96e-05 | 9.43e-05 | -0.420 | 0.675 |
| Time × XBB/Omicron (recombinant) | 3.49e-05 | 1.09e-04 | 0.321 | 0.748 |
| Time × Other | 8.87e-04 | 7.74e-05 | 11.500 | <0.001 |

## Appendix

### The COVID-19 Genomics UK (COG-UK) consortium FINAL (06-2021 V4)

**Funding acquisition, Leadership and supervision, Metadata curation, Project administration, Samples and logistics, Sequencing and analysis, Software and analysis tools, and Visualisation:**

Dr Samuel C Robson PhD ^13, 84^

**Funding acquisition, Leadership and supervision, Metadata curation, Project administration, Samples and logistics, Sequencing and analysis, and Software and analysis tools:**

Dr Thomas R Connor PhD ^11, 74^ and Prof Nicholas J Loman PhD ^43^

**Leadership and supervision, Metadata curation, Project administration, Samples and logistics, Sequencing and analysis, Software and analysis tools, and Visualisation:**

Dr Tanya Golubchik PhD 5

**Funding acquisition, Leadership and supervision, Metadata curation, Samples and logistics, Sequencing and analysis, and Visualisation:**

Dr Rocio T Martinez Nunez PhD ^46^

**Funding acquisition, Leadership and supervision, Project administration, Samples and logistics, Sequencing and analysis, and Software and analysis tools:**

Dr David Bonsall PhD ^5^

**Funding acquisition, Leadership and supervision, Project administration, Sequencing and analysis, Software and analysis tools, and Visualisation:**

Prof Andrew Rambaut DPhil ^104^

**Funding acquisition, Metadata curation, Project administration, Samples and logistics, Sequencing and analysis, and Software and analysis tools:**

Dr Luke B Snell MSc, MBBS ^12^

**Leadership and supervision, Metadata curation, Project administration, Samples and logistics, Software and analysis tools, and Visualisation:**

Rich Livett MSc ^116^

**Funding acquisition, Leadership and supervision, Metadata curation, Project administration, and Samples and logistics:**

Dr Catherine Ludden PhD ^20, 70^

**Funding acquisition, Leadership and supervision, Metadata curation, Samples and logistics, and Sequencing and analysis:**

Dr Sally Corden PhD ^74^ and Dr Eleni Nastouli FRCPath ^96, 95, 30^

**Funding acquisition, Leadership and supervision, Metadata curation, Sequencing and analysis, and Software and analysis tools:**

Dr Gaia Nebbia PhD, FRCPath ^12^

**Funding acquisition, Leadership and supervision, Project administration, Samples and logistics, and Sequencing and analysis:**

Ian Johnston BSc ^116^

**Leadership and supervision, Metadata curation, Project administration, Samples and logistics, and Sequencing and analysis:**

Prof Katrina Lythgoe PhD ^5^, Dr M. Estee Torok FRCP ^19, 20^ and Prof Ian G Goodfellow PhD ^24^

**Leadership and supervision, Metadata curation, Project administration, Samples and logistics, and Visualisation:**

Dr Jacqui A Prieto PhD ^97, 82^ and Dr Kordo Saeed MD, FRCPath ^97, 83^

**Leadership and supervision, Metadata curation, Project administration, Sequencing and analysis, and Software and analysis tools:**

Dr David K Jackson PhD ^116^

**Leadership and supervision, Metadata curation, Samples and logistics, Sequencing and analysis, and Visualisation:**

Dr Catherine Houlihan PhD ^96, 94^

**Leadership and supervision, Metadata curation, Sequencing and analysis, Software and analysis tools, and Visualisation:**

Dr Dan Frampton PhD ^94, 95^

**Metadata curation, Project administration, Samples and logistics, Sequencing and analysis, and Software and analysis tools:**

Dr William L Hamilton PhD ^19^ and Dr Adam A Witney PhD ^41^

**Funding acquisition, Samples and logistics, Sequencing and analysis, and Visualisation:**

Dr Giselda Bucca PhD ^101^

**Funding acquisition, Leadership and supervision, Metadata curation, and Project administration:**

Dr Cassie F Pope PhD^40, 41^

**Funding acquisition, Leadership and supervision, Metadata curation, and Samples and logistics:**

Dr Catherine Moore PhD ^74^

**Funding acquisition, Leadership and supervision, Metadata curation, and Sequencing and analysis:**

Prof Emma C Thomson PhD, FRCP ^53^

**Funding acquisition, Leadership and supervision, Project administration, and Samples and logistics:**

Dr Teresa Cutino-Moguel PhD ^2^, Dr Ewan M Harrison PhD ^116, 102^

**Funding acquisition, Leadership and supervision, Sequencing and analysis, and Visualisation:**

Prof Colin P Smith PhD ^101^

**Leadership and supervision, Metadata curation, Project administration, and Sequencing and analysis:**

Fiona Rogan BSc ^77^

**Leadership and supervision, Metadata curation, Project administration, and Samples and logistics:**

Shaun M Beckwith MSc ^6^, Abigail Murray Degree ^6^, Dawn Singleton HNC ^6^, Dr Kirstine Eastick PhD, FRCPath ^37^, Dr Liz A Sheridan PhD ^98^, Paul Randell MSc, PgD ^99^, Dr Leigh M Jackson PhD ^105^, Dr Cristina V Ariani PhD ^116^ and Dr Sónia Gonçalves PhD ^116^

**Leadership and supervision, Metadata curation, Samples and logistics, and Sequencing and analysis:**

Dr Derek J Fairley PhD ^3, 77^, Prof Matthew W Loose PhD ^18^ and Joanne Watkins MSc ^74^

**Leadership and supervision, Metadata curation, Samples and logistics, and Visualisation:**

Dr Samuel Moses MD ^25, 106^

**Leadership and supervision, Metadata curation, Sequencing and analysis, and Software and analysis tools:**

Dr Sam Nicholls PhD ^43^, Dr Matthew Bull PhD ^74^ and Dr Roberto Amato PhD ^116^

**Leadership and supervision, Project administration, Samples and logistics, and Sequencing and analysis:**

Prof Darren L Smith PhD ^36, 65, 66^

**Leadership and supervision, Sequencing and analysis, Software and analysis tools, and Visualisation:**

Prof David M Aanensen PhD ^14, 116^ and Dr Jeffrey C Barrett PhD ^116^

**Metadata curation, Project administration, Samples and logistics, and Sequencing and analysis:**

Dr Beatrix Kele PhD ^2^, Dr Dinesh Aggarwal MRCP^20, 116, 70^, Dr James G Shepherd MBCHB, MRCP ^53^, Dr Martin D Curran PhD ^71^ and Dr Surendra Parmar PhD ^71^

**Metadata curation, Project administration, Sequencing and analysis, and Software and analysis tools:**

Dr Matthew D Parker PhD ^109^

**Metadata curation, Samples and logistics, Sequencing and analysis, and Software and analysis tools:**

Dr Catryn Williams PhD ^74^

**Metadata curation, Samples and logistics, Sequencing and analysis, and Visualisation:**

Dr Sharon Glaysher PhD ^68^

**Metadata curation, Sequencing and analysis, Software and analysis tools, and Visualisation:**

Dr Anthony P Underwood PhD ^14, 116^, Dr Matthew Bashton PhD ^36, 65^, Dr Nicole Pacchiarini PhD ^74^, Dr Katie F Loveson PhD ^84^ and Matthew Byott MSc ^95, 96^

**Project administration, Sequencing and analysis, Software and analysis tools, and Visualisation:**

Dr Alessandro M Carabelli PhD ^20^

**Funding acquisition, Leadership and supervision, and Metadata curation:**

Dr Kate E Templeton PhD ^56, 104^

**Funding acquisition, Leadership and supervision, and Project administration:**

Prof Sharon J Peacock PhD ^20, 70^, Dr Thushan I de Silva PhD ^109^, Dr Dennis Wang PhD ^109^, Dr Cordelia F Langford PhD ^116^ and John Sillitoe BEng ^116^

**Funding acquisition, Leadership and supervision, and Samples and logistics:**

Prof Rory N Gunson PhD, FRCPath ^55^

**Funding acquisition, Leadership and supervision, and Sequencing and analysis:**

Dr Simon Cottrell PhD ^74^, Dr Justin O’Grady PhD ^75, 103^ and Prof Dominic Kwiatkowski PhD^116, 108^

**Leadership and supervision, Metadata curation, and Project administration:**

Dr Patrick J Lillie PhD, FRCP ^37^

**Leadership and supervision, Metadata curation, and Samples and logistics:**

Dr Nicholas Cortes MBCHB ^33^, Dr Nathan Moore MBCHB ^33^, Dr Claire Thomas DPhil ^33^, Phillipa J Burns MSc, DipRCPath ^37^, Dr Tabitha W Mahungu FRCPath ^80^ and Steven Liggett BSc ^86^

**Leadership and supervision, Metadata curation, and Sequencing and analysis:**

Angela H Beckett MSc ^13, 81^ and Prof Matthew TG Holden PhD ^73^

**Leadership and supervision, Project administration, and Samples and logistics:**

Dr Lisa J Levett PhD ^34^, Dr Husam Osman PhD ^70, 35^ and Dr Mohammed O Hassan-Ibrahim PhD, FRCPath ^99^

**Leadership and supervision, Project administration, and Sequencing and analysis:**

Dr David A Simpson PhD ^77^

**Leadership and supervision, Samples and logistics, and Sequencing and analysis:** Dr Meera Chand PhD ^72^, Prof Ravi K Gupta PhD ^102^, Prof Alistair C Darby PhD ^107^ and Prof Steve Paterson PhD ^107^

**Leadership and supervision, Sequencing and analysis, and Software and analysis tools:**

Prof Oliver G Pybus DPhil ^23^, Dr Erik M Volz PhD ^39^, Prof Daniela de Angelis PhD ^52^, Prof David L Robertson PhD ^53^, Dr Andrew J Page PhD ^75^ and Dr Inigo Martincorena PhD ^116^

**Leadership and supervision, Sequencing and analysis, and Visualisation:**

Dr Louise Aigrain PhD ^116^ and Dr Andrew R Bassett PhD ^116^

**Metadata curation, Project administration, and Samples and logistics:**

Dr Nick Wong DPhil, MRCP, FRCPath ^50^, Dr Yusri Taha MD, PhD ^89^, Michelle J Erkiert BA ^99^ and Dr Michael H Spencer Chapman MBBS ^116, 102^

**Metadata curation, Project administration, and Sequencing and analysis:**

Dr Rebecca Dewar PhD ^56^ and Martin P McHugh MSc ^56, 111^

**Metadata curation, Project administration, and Software and analysis tools:**

Siddharth Mookerjee MPH ^38, 57^

**Metadata curation, Project administration, and Visualisation:**

Stephen Aplin ^97^, Matthew Harvey ^97^, Thea Sass ^97^, Dr Helen Umpleby FRCP ^97^ and Helen Wheeler ^97^

**Metadata curation, Samples and logistics, and Sequencing and analysis:**

Dr James P McKenna PhD ^3^, Dr Ben Warne MRCP ^9^, Joshua F Taylor MSc ^22^, Yasmin Chaudhry BSc ^24^, Rhys Izuagbe ^24^, Dr Aminu S Jahun PhD ^24^, Dr Gregory R Young PhD ^36, 65^, Dr Claire McMurray PhD ^43^, Dr Clare M McCann PhD ^65, 66^, Dr Andrew Nelson PhD ^65, 66^ and Scott Elliott ^68^

**Metadata curation, Samples and logistics, and Visualisation:**

Hannah Lowe MSc ^25^

**Metadata curation, Sequencing and analysis, and Software and analysis tools:**

Dr Anna Price PhD ^11^, Matthew R Crown BSc ^65^, Dr Sara Rey PhD ^74^, Dr Sunando Roy PhD^96^ and Dr Ben Temperton PhD ^105^

**Metadata curation, Sequencing and analysis, and Visualisation:**

Dr Sharif Shaaban PhD ^73^ and Dr Andrew R Hesketh PhD ^101^

**Project administration, Samples and logistics, and Sequencing and analysis:**

Dr Kenneth G Laing PhD^41^, Dr Irene M Monahan PhD ^41^ and Dr Judith Heaney PhD ^95, 96, 34^

**Project administration, Samples and logistics, and Visualisation:**

Dr Emanuela Pelosi FRCPath ^97^, Siona Silviera MSc ^97^ and Dr Eleri Wilson-Davies MD, FRCPath ^97^

**Samples and logistics, Software and analysis tools, and Visualisation:**

Dr Helen Fryer PhD ^5^

**Sequencing and analysis, Software and analysis tools, and Visualization:**

Dr Helen Adams PhD ^4^, Dr Louis du Plessis PhD ^23^, Dr Rob Johnson PhD ^39^, Dr William T Harvey PhD ^53, 42^, Dr Joseph Hughes PhD ^53^, Dr Richard J Orton PhD ^53^, Dr Lewis G Spurgin PhD ^59^, Dr Yann Bourgeois PhD ^81^, Dr Chris Ruis PhD ^102^, Áine O’Toole MSc ^104^, Marina Gourtovaia MSc ^116^ and Dr Theo Sanderson PhD ^116^

**Funding acquisition, and Leadership and supervision:**

Dr Christophe Fraser PhD ^5^, Dr Jonathan Edgeworth PhD, FRCPath ^12^, Prof Judith Breuer MD ^96, 29^, Dr Stephen L Michell PhD ^105^ and Prof John A Todd PhD ^115^

**Funding acquisition, and Project administration:**

Michaela John BSc ^10^ and Dr David Buck PhD ^115^

**Leadership and supervision, and Metadata curation:**

Dr Kavitha Gajee MBBS, FRCPath ^37^ and Dr Gemma L Kay PhD ^75^

**Leadership and supervision, and Project administration:**

David Heyburn ^74^

**Leadership and supervision, and Samples and logistics:**

Dr Themoula Charalampous PhD ^12, 46^, Adela Alcolea-Medina ^32, 112^, Katie Kitchman BSc ^37^, Prof Alan McNally PhD ^43, 93^, David T Pritchard MSc, CSci ^50^, Dr Samir Dervisevic FRCPath ^58^, Dr Peter Muir PhD ^70^, Dr Esther Robinson PhD ^70, 35^, Dr Barry B Vipond PhD ^70^, Newara A Ramadan MSc, CSci, FIBMS ^78^, Dr Christopher Jeanes MBBS ^90^, Danni Weldon BSc ^116^, Jana Catalan MSc ^118^ and Neil Jones MSc ^118^

**Leadership and supervision, and Sequencing and analysis:**

Dr Ana da Silva Filipe PhD ^53^, Dr Chris Williams MBBS ^74^, Marc Fuchs BSc ^77^, Dr Julia Miskelly PhD ^77^, Dr Aaron R Jeffries PhD ^105^, Karen Oliver BSc ^116^ and Dr Naomi R Park PhD^116^

**Metadata curation, and Samples and logistics:**

Amy Ash BSc ^1^, Cherian Koshy MSc, CSci, FIBMS ^1^, Magdalena Barrow ^7^, Dr Sarah L Buchan PhD ^7^, Dr Anna Mantzouratou PhD ^7^, Dr Gemma Clark PhD ^15^, Dr Christopher W Holmes PhD ^16^, Sharon Campbell MSc ^17^, Thomas Davis MSc ^21^, Ngee Keong Tan MSc ^22^, Dr Julianne R Brown PhD ^29^, Dr Kathryn A Harris PhD ^29, 2^, Stephen P Kidd MSc ^33^, Dr Paul R Grant PhD ^34^, Dr Li Xu-McCrae PhD ^35^, Dr Alison Cox PhD ^38, 63^, Pinglawathee Madona^38, 63^, Dr Marcus Pond PhD ^38, 63^, Dr Paul A Randell MBBCh ^38, 63^, Karen T Withell FIBMS ^48^, Cheryl Williams MSc ^51^, Dr Clive Graham MD ^60^, Rebecca Denton-Smith BSc ^62^, Emma Swindells BSc ^62^, Robyn Turnbull BSc ^62^, Dr Tim J Sloan PhD ^67^, Dr Andrew Bosworth PhD ^70, 35^, Stephanie Hutchings ^70^, Hannah M Pymont MSc ^70^, Dr Anna Casey PhD ^76^, Dr Liz Ratcliffe PhD ^76^, Dr Christopher R Jones PhD ^79, 105^, Dr Bridget A Knight PhD ^79, 105^, Dr Tanzina Haque PhD, FRCPath ^80^, Dr Jennifer Hart MRCP ^80^, Dr Dianne Irish-Tavares FRCPath ^80^, Eric Witele MSc ^80^, Craig Mower BA ^86^, Louisa K Watson DipHE ^86^, Jennifer Collins BSc ^89^, Gary Eltringham BSc ^89^, Dorian Crudgington ^98^, Ben Macklin ^98^, Prof Miren Iturriza-Gomara PhD ^107^, Dr Anita O Lucaci PhD ^107^ and Dr Patrick C McClure PhD ^113^

**Metadata curation, and Sequencing and analysis:**

Matthew Carlile BSc ^18^, Dr Nadine Holmes PhD ^18^, Dr Christopher Moore PhD ^18^, Dr Nathaniel Storey PhD ^29^, Dr Stefan Rooke PhD ^73^, Dr Gonzalo Yebra PhD ^73^, Dr Noel Craine DPhil ^74^, Malorie Perry MSc ^74^, Dr Nabil-Fareed Alikhan PhD ^75^, Dr Stephen Bridgett PhD ^77^, Kate F Cook MScR ^84^, Christopher Fearn MSc ^84^, Dr Salman Goudarzi PhD ^84^, Prof Ronan A Lyons MD ^88^, Dr Thomas Williams MD ^104^, Dr Sam T Haldenby PhD ^107^, Jillian Durham BSc^116^ and Dr Steven Leonard PhD ^116^

**Metadata curation, and Software and analysis tools:**

Robert M Davies MA (Cantab) ^116^

**Project administration, and Samples and logistics:**

Dr Rahul Batra MD ^12^, Beth Blane BSc ^20^, Dr Moira J Spyer PhD ^30, 95, 96^, Perminder Smith MSc ^32, 112^, Mehmet Yavus ^85, 109^, Dr Rachel J Williams PhD ^96^, Dr Adhyana IK Mahanama MD ^97^, Dr Buddhini Samaraweera MD ^97^, Sophia T Girgis MSc ^102^, Samantha E Hansford CSci ^109^, Dr Angie Green PhD ^115^, Dr Charlotte Beaver PhD ^116^, Katherine L Bellis ^116, 102^, Matthew J Dorman ^116^, Sally Kay ^116^, Liam Prestwood ^116^ and Dr Shavanthi Rajatileka PhD^116^

**Project administration, and Sequencing and analysis:**

Dr Joshua Quick PhD ^43^

**Project administration, and Software and analysis tools:**

Radoslaw Poplawski BSc ^43^

**Samples and logistics, and Sequencing and analysis:**

Dr Nicola Reynolds PhD ^8^, Andrew Mack MPhil ^11^, Dr Arthur Morriss PhD ^11^, Thomas Whalley BSc ^11^, Bindi Patel BSc ^12^, Dr Iliana Georgana PhD ^24^, Dr Myra Hosmillo PhD ^24^, Malte L Pinckert MPhil ^24^, Dr Joanne Stockton PhD ^43^, Dr John H Henderson PhD ^65^, Amy Hollis HND ^65^, Dr William Stanley PhD ^65^, Dr Wen C Yew PhD ^65^, Dr Richard Myers PhD ^72^, Dr Alicia Thornton PhD ^72^, Alexander Adams BSc ^74^, Tara Annett BSc ^74^, Dr Hibo Asad PhD ^74^, Alec Birchley MSc ^74^, Jason Coombes BSc ^74^, Johnathan M Evans MSc ^74^, Laia Fina ^74^, Bree Gatica-Wilcox MPhil ^74^, Lauren Gilbert ^74^, Lee Graham BSc ^74^, Jessica Hey BSc ^74^, Ember Hilvers MPH ^74^, Sophie Jones MSc ^74^, Hannah Jones ^74^, Sara Kumziene-Summerhayes MSc ^74^, Dr Caoimhe McKerr PhD ^74^, Jessica Powell BSc ^74^, Georgia Pugh ^74^, Sarah Taylor ^74^, Alexander J Trotter MRes ^75^, Charlotte A Williams BSc ^96^, Leanne M Kermack MSc ^102^, Benjamin H Foulkes MSc ^109^, Marta Gallis MSc ^109^, Hailey R Hornsby MSc ^109^, Stavroula F Louka MSc ^109^, Dr Manoj Pohare PhD ^109^, Paige Wolverson MSc ^109^, Peijun Zhang MSc ^109^, George MacIntyre-Cockett BSc ^115^, Amy Trebes MSc ^115^, Dr Robin J Moll PhD ^116^, Lynne Ferguson MSc ^117^, Dr Emily J Goldstein PhD ^117^, Dr Alasdair Maclean PhD ^117^ and Dr Rachael Tomb PhD ^117^

**Samples and logistics, and Software and analysis tools:**

Dr Igor Starinskij MSc, MRCP ^53^

**Sequencing and analysis, and Software and analysis tools:**

Laura Thomson BSc ^5^, Joel Southgate MSc ^11, 74^, Dr Moritz UG Kraemer DPhil ^23^, Dr Jayna Raghwani PhD ^23^, Dr Alex E Zarebski PhD ^23^, Olivia Boyd MSc ^39^, Lily Geidelberg MSc ^39^, Dr Chris J Illingworth PhD ^52^, Dr Chris Jackson PhD ^52^, Dr David Pascall PhD ^52^, Dr Sreenu Vattipally PhD ^53^, Timothy M Freeman MPhil ^109^, Dr Sharon N Hsu PhD ^109^, Dr Benjamin B Lindsey MRCP ^109^, Dr Keith James PhD ^116^, Kevin Lewis ^116^, Gerry Tonkin-Hill ^116^ and Dr Jaime M Tovar-Corona PhD ^116^

**Sequencing and analysis, and Visualisation:**

MacGregor Cox MSci ^20^

**Software and analysis tools, and Visualisation:**

Dr Khalil Abudahab PhD ^14, 116^, Mirko Menegazzo ^14^, Ben EW Taylor MEng ^14, 116^, Dr Corin A Yeats PhD ^14^, Afrida Mukaddas BTech ^53^, Derek W Wright MSc ^53^, Dr Leonardo de Oliveira Martins PhD ^75^, Dr Rachel Colquhoun DPhil ^104^, Verity Hill ^104^, Dr Ben Jackson PhD ^104^, Dr JT McCrone PhD ^104^, Dr Nathan Medd PhD ^104^, Dr Emily Scher PhD ^104^ and Jon-Paul Keatley ^116^

**Leadership and supervision:**

Dr Tanya Curran PhD ^3^, Dr Sian Morgan FRCPath ^10^, Prof Patrick Maxwell PhD ^20^, Prof Ken Smith PhD ^20^, Dr Sahar Eldirdiri MBBS, MSc, FRCPath ^21^, Anita Kenyon MSc ^21^, Prof Alison H Holmes MD ^38, 57^, Dr James R Price PhD ^38, 57^, Dr Tim Wyatt PhD ^69^, Dr Alison E Mather PhD ^75^, Dr Timofey Skvortsov PhD ^77^ and Prof John A Hartley PhD ^96^

**Metadata curation:**

Prof Martyn Guest PhD ^11^, Dr Christine Kitchen PhD ^11^, Dr Ian Merrick PhD ^11^, Robert Munn BSc ^11^, Dr Beatrice Bertolusso Degree ^33^, Dr Jessica Lynch MBCHB ^33^, Dr Gabrielle Vernet MBBS ^33^, Stuart Kirk MSc ^34^, Dr Elizabeth Wastnedge MD ^56^, Dr Rachael Stanley PhD ^58^, Giles Idle ^64^, Dr Declan T Bradley PhD ^69, 77^, Nicholas F Killough MSc ^69^, Dr Jennifer Poyner MD ^79^ and Matilde Mori BSc ^110^

**Project administration:**

Owen Jones BSc ^11^, Victoria Wright BSc ^18^, Ellena Brooks MA ^20^, Carol M Churcher BSc ^20^, Dr Laia Delgado Callico PhD ^20^, Mireille Fragakis HND ^20^, Dr Katerina Galai PhD ^20, 70^, Dr Andrew Jermy PhD ^20^, Sarah Judges BA ^20^, Anna Markov BSc ^20^, Georgina M McManus BSc ^20^, Kim S Smith ^20^, Peter M D Thomas-McEwen MSc ^20^, Dr Elaine Westwick PhD ^20^, Dr Stephen W Attwood PhD ^23^, Dr Frances Bolt PhD ^38, 57^, Dr Alisha Davies PhD ^74^, Elen De Lacy MPH ^74^, Fatima Downing ^74^, Sue Edwards ^74^, Lizzie Meadows MA ^75^, Sarah Jeremiah MSc ^97^, Dr Nikki Smith PhD ^109^ and Luke Foulser ^116^

**Samples and logistics:**

Amita Patel BSc ^12^, Dr Louise Berry PhD ^15^, Dr Tim Boswell PhD ^15^, Dr Vicki M Fleming PhD ^15^, Dr Hannah C Howson-Wells PhD ^15^, Dr Amelia Joseph PhD ^15^, Manjinder Khakh ^15^, Dr Michelle M Lister PhD ^15^, Paul W Bird MSc, MRes ^16^, Karlie Fallon ^16^, Thomas Helmer ^16^, Dr Claire L McMurray PhD ^16^, Mina Odedra BSc ^16^, Jessica Shaw BSc ^16^, Dr Julian W Tang PhD ^16^, Nicholas J Willford MSc ^16^, Victoria Blakey BSc ^17^, Dr Veena Raviprakash MD ^17^, Nicola Sheriff BSc ^17^, Lesley-Anne Williams BSc ^17^, Theresa Feltwell MSc ^20^, Dr Luke Bedford PhD ^26^, Dr James S Cargill PhD ^27^, Warwick Hughes MSc ^27^, Dr Jonathan Moore MD ^28^, Susanne Stonehouse BSc ^28^, Laura Atkinson MSc ^29^, Jack CD Lee MSc ^29^, Dr Divya Shah PhD ^29^, Natasha Ohemeng-Kumi MSc ^32, 112^, John Ramble MSc ^32, 112^, Jasveen Sehmi MSc ^32, 112^, Dr Rebecca Williams BMBS ^33^, Wendy Chatterton MSc ^34^, Monika Pusok MSc ^34^, William Everson MSc ^37^, Anibolina Castigador IBMS HCPC ^44^, Emily Macnaughton FRCPath ^44^, Dr Kate El Bouzidi MRCP ^45^, Dr Temi Lampejo FRCPath ^45^, Dr Malur Sudhanva FRCPath ^45^, Cassie Breen BSc ^47^, Dr Graciela Sluga MD, MSc ^48^, Dr Shazaad SY Ahmad MSc ^49, 70^, Dr Ryan P George PhD ^49^, Dr Nicholas W Machin MSc ^49, 70^, Debbie Binns BSc ^50^, Victoria James BSc ^50^, Dr Rachel Blacow MBCHB ^55^, Dr Lindsay Coupland PhD ^58^, Dr Louise Smith PhD ^59^, Dr Edward Barton MD ^60^, Debra Padgett BSc ^60^, Garren Scott BSc ^60^, Dr Aidan Cross MBCHB ^61^, Dr Mariyam Mirfenderesky FRCPath ^61^, Jane Greenaway MSc ^62^, Kevin Cole ^64^, Phillip Clarke ^67^, Nichola Duckworth ^67^, Sarah Walsh ^67^, Kelly Bicknell ^68^, Robert Impey MSc ^68^, Dr Sarah Wyllie PhD ^68^, Richard Hopes ^70^, Dr Chloe Bishop PhD ^72^, Dr Vicki Chalker PhD ^72^, Dr Ian Harrison PhD ^72^, Laura Gifford MSc ^74^, Dr Zoltan Molnar PhD ^77^, Dr Cressida Auckland FRCPath ^79^, Dr Cariad Evans PhD ^85, 109^, Dr Kate Johnson PhD ^85, 109^, Dr David G Partridge FRCP, FRCPath ^85, 109^, Dr Mohammad Raza PhD ^85, 109^, Paul Baker MD ^86^, Prof Stephen Bonner PhD ^86^, Sarah Essex ^86^, Leanne J Murray ^86^, Andrew I Lawton MSc ^87^, Dr Shirelle Burton-Fanning MD ^89^, Dr Brendan AI Payne MD ^89^, Dr Sheila Waugh MD ^89^, Andrea N Gomes MSc ^91^, Maimuna Kimuli MSc ^91^, Darren R Murray MSc ^91^, Paula Ashfield MSc ^92^, Dr Donald Dobie MBCHB ^92^, Dr Fiona Ashford PhD ^93^, Dr Angus Best PhD ^93^, Dr Liam Crawford PhD ^93^, Dr Nicola Cumley PhD ^93^, Dr Megan Mayhew PhD ^93^, Dr Oliver Megram PhD ^93^, Dr Jeremy Mirza PhD ^93^, Dr Emma Moles-Garcia PhD ^93^, Dr Benita Percival PhD ^93^, Megan Driscoll BSc ^96^, Leah Ensell BSc ^96^, Dr Helen L Lowe PhD ^96^, Laurentiu Maftei BSc ^96^, Matteo Mondani MSc ^96^, Nicola J Chaloner BSc ^99^, Benjamin J Cogger BSc ^99^, Lisa J Easton MSc ^99^, Hannah Huckson BSc ^99^, Jonathan Lewis MSc, PgD, FIBMS ^99^, Sarah Lowdon BSc ^99^, Cassandra S Malone MSc ^99^, Florence Munemo BSc ^99^, Manasa Mutingwende MSc ^99^, Roberto Nicodemi BSc ^99^, Olga Podplomyk FD ^99^, Thomas Somassa BSc ^99^, Dr Andrew Beggs PhD ^100^, Dr Alex Richter PhD ^100^, Claire Cormie ^102^, Joana Dias MSc ^102^, Sally Forrest BSc ^102^, Dr Ellen E Higginson PhD ^102^, Mailis Maes MPhil ^102^, Jamie Young BSc ^102^, Dr Rose K Davidson PhD ^103^, Kathryn A Jackson MSc ^107^, Dr Alexander J Keeley MRCP ^109^, Prof Jonathan Ball PhD ^113^, Timothy Byaruhanga MSc ^113^, Dr Joseph G Chappell PhD ^113^, Jayasree Dey MSc ^113^, Jack D Hill MSc ^113^, Emily J Park MSc ^113^, Arezou Fanaie MSc ^114^, Rachel A Hilson MSc ^114^, Geraldine Yaze MSc ^114^ and Stephanie Lo ^116^

**Sequencing and analysis:**

Safiah Afifi BSc ^10^, Robert Beer BSc ^10^, Joshua Maksimovic FD ^10^, Kathryn McCluggage Masters ^10^, Karla Spellman FD ^10^, Catherine Bresner BSc ^11^, William Fuller BSc ^11^, Dr Angela Marchbank BSc ^11^, Trudy Workman HNC ^11^, Dr Ekaterina Shelest PhD ^13, 81^, Dr Johnny Debebe PhD ^18^, Dr Fei Sang PhD ^18^, Dr Sarah Francois PhD ^23^, Bernardo Gutierrez MSc ^23^, Dr Tetyana I Vasylyeva DPhil ^23^, Dr Flavia Flaviani PhD ^31^, Dr Manon Ragonnet-Cronin PhD ^39^, Dr Katherine L Smollett PhD ^42^, Alice Broos BSc ^53^, Daniel Mair BSc ^53^, Jenna Nichols BSc ^53^, Dr Kyriaki Nomikou PhD ^53^, Dr Lily Tong PhD ^53^, Ioulia Tsatsani MSc ^53^, Prof Sarah O’Brien PhD ^54^, Prof Steven Rushton PhD ^54^, Dr Roy Sanderson PhD ^54^, Dr Jon Perkins MBCHB ^55^, Seb Cotton MSc ^56^, Abbie Gallagher BSc ^56^, Dr Elias Allara MD, PhD ^70, 102^, Clare Pearson MSc ^70, 102^, Dr David Bibby PhD ^72^, Dr Gavin Dabrera PhD ^72^, Dr Nicholas Ellaby PhD ^72^, Dr Eileen Gallagher PhD ^72^, Dr Jonathan Hubb PhD ^72^, Dr Angie Lackenby PhD ^72^, Dr David Lee PhD ^72^, Nikos Manesis ^72^, Dr Tamyo Mbisa PhD ^72^, Dr Steven Platt PhD ^72^, Katherine A Twohig ^72^, Dr Mari Morgan PhD ^74^, Alp Aydin MSci ^75^, David J Baker BEng ^75^, Dr Ebenezer Foster-Nyarko PhD ^75^, Dr Sophie J Prosolek PhD ^75^, Steven Rudder ^75^, Chris Baxter BSc ^77^, Sílvia F Carvalho MSc ^77^, Dr Deborah Lavin PhD ^77^, Dr Arun Mariappan PhD ^77^, Dr Clara Radulescu PhD ^77^, Dr Aditi Singh PhD ^77^, Miao Tang MD ^77^, Helen Morcrette BSc ^79^, Nadua Bayzid BSc ^96^, Marius Cotic MSc ^96^, Dr Carlos E Balcazar PhD ^104^, Dr Michael D Gallagher PhD ^104^, Dr Daniel Maloney PhD ^104^, Thomas D Stanton BSc ^104^, Dr Kathleen A Williamson PhD ^104^, Dr Robin Manley PhD ^105^, Michelle L Michelsen BSc ^105^, Dr Christine M Sambles PhD ^105^, Dr David J Studholme PhD ^105^, Joanna Warwick-Dugdale BSc ^105^, Richard Eccles MSc ^107^, Matthew Gemmell MSc ^107^, Dr Richard Gregory PhD ^107^, Dr Margaret Hughes PhD ^107^, Charlotte Nelson MSc ^107^, Dr Lucille Rainbow PhD ^107^, Dr Edith E Vamos PhD ^107^, Hermione J Webster BSc ^107^, Dr Mark Whitehead PhD ^107^, Claudia Wierzbicki BSc ^107^, Dr Adrienn Angyal PhD ^109^, Dr Luke R Green PhD ^109^, Dr Max Whiteley PhD ^109^, Emma Betteridge BSc ^116^, Dr Iraad F Bronner PhD ^116^, Ben W Farr BSc ^116^, Scott Goodwin MSc ^116^, Dr Stefanie V Lensing PhD ^116^, Shane A McCarthy ^116, 102^, Dr Michael A Quail PhD ^116^, Diana Rajan MSc ^116^, Dr Nicholas M Redshaw PhD ^116^, Carol Scott ^116^, Lesley Shirley MSc ^116^ and Scott AJ Thurston BSc ^116^

**Software and analysis tools:**

Dr Will Rowe PhD^43^, Amy Gaskin MSc ^74^, Dr Thanh Le-Viet PhD ^75^, James Bonfield BSc ^116^, Jennifier Liddle ^116^ and Andrew Whitwham BSc ^116^

**1** Barking, Havering and Redbridge University Hospitals NHS Trust, **2** Barts Health NHS Trust, **3** Belfast Health & Social Care Trust, **4** Betsi Cadwaladr University Health Board, **5** Big Data Institute, Nuffield Department of Medicine, University of Oxford, **6** Blackpool Teaching Hospitals NHS Foundation Trust, **7** Bournemouth University, **8** Cambridge Stem Cell Institute, University of Cambridge, **9** Cambridge University Hospitals NHS Foundation Trust, **10** Cardiff and Vale University Health Board, **11** Cardiff University, **12** Centre for Clinical Infection and Diagnostics Research, Department of Infectious Diseases, Guy’s and St Thomas’ NHS Foundation Trust, **13** Centre for Enzyme Innovation, University of Portsmouth, **14** Centre for Genomic Pathogen Surveillance, University of Oxford, **15** Clinical Microbiology Department, Queens Medical Centre, Nottingham University Hospitals NHS Trust, **16** Clinical Microbiology, University Hospitals of Leicester NHS Trust, **17** County Durham and Darlington NHS Foundation Trust, **18** Deep Seq, School of Life Sciences, Queens Medical Centre, University of Nottingham, **19** Department of Infectious Diseases and Microbiology, Cambridge University Hospitals NHS Foundation Trust, **20** Department of Medicine, University of Cambridge, **21** Department of Microbiology, Kettering General Hospital, **22** Department of Microbiology, South West London Pathology, **23** Department of Zoology, University of Oxford, **24** Division of Virology, Department of Pathology, University of Cambridge, **25** East Kent Hospitals University NHS Foundation Trust, **26** East Suffolk and North Essex NHS Foundation Trust, **27** East Sussex Healthcare NHS Trust**, 28** Gateshead Health NHS Foundation Trust, **29** Great Ormond Street Hospital for Children NHS Foundation Trust, **30** Great Ormond Street Institute of Child Health (GOS ICH), University College London (UCL), **31** Guy’s and St. Thomas’ Biomedical Research Centre, **32** Guy’s and St. Thomas’ NHS Foundation Trust, **33** Hampshire Hospitals NHS Foundation Trust, **34** Health Services Laboratories, **35** Heartlands Hospital, Birmingham, **36** Hub for Biotechnology in the Built Environment, Northumbria University, **37** Hull University Teaching Hospitals NHS Trust, **38** Imperial College Healthcare NHS Trust, **39** Imperial College London, **40** Infection Care Group, St George’s University Hospitals NHS Foundation Trust, **41** Institute for Infection and Immunity, St George’s University of London, **42** Institute of Biodiversity, Animal Health & Comparative Medicine, **43** Institute of Microbiology and Infection, University of Birmingham, **44** Isle of Wight NHS Trust, **45** King’s College Hospital NHS Foundation Trust, **46** King’s College London, **47** Liverpool Clinical Laboratories, **48** Maidstone and Tunbridge Wells NHS Trust, **49** Manchester University NHS Foundation Trust, **50** Microbiology Department, Buckinghamshire Healthcare NHS Trust, **51** Microbiology, Royal Oldham Hospital, **52** MRC Biostatistics Unit, University of Cambridge, **53** MRC-University of Glasgow Centre for Virus Research, **54** Newcastle University, **55** NHS Greater Glasgow and Clyde, **56** NHS Lothian, **57** NIHR Health Protection Research Unit in HCAI and AMR, Imperial College London, **58** Norfolk and Norwich University Hospitals NHS Foundation Trust, **59** Norfolk County Council, **60** North Cumbria Integrated Care NHS Foundation Trust, **61** North Middlesex University Hospital NHS Trust, **62** North Tees and Hartlepool NHS Foundation Trust, **63** North West London Pathology, **64** Northumbria Healthcare NHS Foundation Trust, **65** Northumbria University, **66** NU-OMICS, Northumbria University, **67** Path Links, Northern Lincolnshire and Goole NHS Foundation Trust, **68** Portsmouth Hospitals University NHS Trust, **69** Public Health Agency, Northern Ireland, **70** Public Health England, **71** Public Health England, Cambridge, **72** Public Health England, Colindale, **73** Public Health Scotland, **74** Public Health Wales, **75** Quadram Institute Bioscience, **76** Queen Elizabeth Hospital, Birmingham, **77** Queen’s University Belfast, **78** Royal Brompton and Harefield Hospitals, **79** Royal Devon and Exeter NHS Foundation Trust, **80** Royal Free London NHS Foundation Trust, **81** School of Biological Sciences, University of Portsmouth, **82** School of Health Sciences, University of Southampton, **83** School of Medicine, University of Southampton, **84** School of Pharmacy & Biomedical Sciences, University of Portsmouth, **85** Sheffield Teaching Hospitals NHS Foundation Trust, **86** South Tees Hospitals NHS Foundation Trust, **87** Southwest Pathology Services, **88** Swansea University, **89** The Newcastle upon Tyne Hospitals NHS Foundation Trust, **90** The Queen Elizabeth Hospital King’s Lynn NHS Foundation Trust, **91** The Royal Marsden NHS Foundation Trust, **92** The Royal Wolverhampton NHS Trust, **93** Turnkey Laboratory, University of Birmingham, **94** University College London Division of Infection and Immunity**, 95** University College London Hospital Advanced Pathogen Diagnostics Unit**, 96** University College London Hospitals NHS Foundation Trust, **97** University Hospital Southampton NHS Foundation Trust, **98** University Hospitals Dorset NHS Foundation Trust, **99** University Hospitals Sussex NHS Foundation Trust, **100** University of Birmingham, **101** University of Brighton, **102** University of Cambridge, **103** University of East Anglia, **104** University of Edinburgh, **105** University of Exeter, **106** University of Kent, **107** University of Liverpool, **108** University of Oxford, **109** University of Sheffield, **110** University of Southampton, **111** University of St Andrews, **112** Viapath, Guy’s and St Thomas’ NHS Foundation Trust, and King’s College Hospital NHS Foundation Trust, **113** Virology, School of Life Sciences, Queens Medical Centre, University of Nottingham, **114** Watford General Hospital, **115** Wellcome Centre for Human Genetics, Nuffield Department of Medicine, University of Oxford, **116** Wellcome Sanger Institute, **117** West of Scotland Specialist Virology Centre, NHS Greater Glasgow and Clyde, **118** Whittington Health NHS Trust

## References

Bonsall, David, Tanya Golubchik, Mariateresa de Cesare, Mohammed Limbada, Barry Kosloff, George MacIntyre-Cockett, Matthew Hall, et al. 2020. “A Comprehensive Genomics Solution for HIV Surveillance and Clinical Monitoring in Low-Income Settings.” Journal of Clinical Microbiology 58 (10). https://doi.org/10.1128/JCM.00382-20.

Carabelli, Alessandro M., Thomas P. Peacock, Lucy G. Thorne, William T. Harvey, Joseph Hughes, Sharon J. Peacock, Wendy S. Barclay, Thushan I. de Silva, Greg J. Towers, and David L. Robertson. 2023. “SARS-CoV-2 Variant Biology: Immune Escape, Transmission and Fitness.” Nature Reviews. Microbiology 21 (3): 162–77.

COG-UK. 2020. “COG-UK Publication.” 2020. https://www.protocols.io/workspaces/coguk/publication.

Davies, Nicholas G., Sam Abbott, Rosanna C. Barnard, Christopher I. Jarvis, Adam J. Kucharski, James D. Munday, Carl A. B. Pearson, et al. 2021. “Estimated Transmissibility and Impact of SARS-CoV-2 Lineage B.1.1.7 in England.” Science 372 (6538). https://doi.org/10.1126/science.abg3055.

Davies, Nicholas G., Christopher I. Jarvis, W. John Edmunds, Nicholas P. Jewell, Karla Diaz-Ordaz, and Ruth H. Keogh. 2021. “Increased Mortality in Community-Tested Cases of SARS-CoV-2 Lineage B.1.1.7.” Nature 593 (7858): 270–74.

Franceschi, Vinícius Bonetti, Andressa Schneiders Santos, Andressa Barreto Glaeser, Janini Cristina Paiz, Gabriel Dickin Caldana, Carem Luana Machado Lessa, Amanda de Menezes Mayer, et al. 2021. “Population-Based Prevalence Surveys during the Covid-19 Pandemic: A Systematic Review.” Reviews in Medical Virology 31 (4): e2200.

Fryer, Helen R., Tanya Golubchik, Matthew Hall, Christophe Fraser, Robert Hinch, Luca Ferretti, Laura Thomson, et al. 2022. “Viral Burdens Are Associated with Age and Viral Variant in a Population-Representative Study of SARS-CoV-2 That Accounts for Time-since-Infection Related Sampling Bias.” bioRxiv. https://doi.org/10.1101/2022.12.02.518847. (PLoS Pathogens, accepted).

Ghafari, Mahan, Matthew Hall, Tanya Golubchik, Daniel Ayoubkhani, Thomas House, George MacIntyre-Cockett, Helen Fryer, et al. 2023. “High Number of SARS-CoV-2 Persistent Infections Uncovered through Genetic Analysis of Samples from a Large Community-Based Surveillance Study.” medRxiv. https://doi.org/10.1101/2023.01.29.23285160.

Golubchik, Tanya, Katrina A. Lythgoe, Matthew Hall, Luca Ferretti, Helen R. Fryer, George MacIntyre-Cockett, Mariateresa de Cesare, et al. 2021. “Early Analysis of a Potential Link between Viral Load and the N501Y Mutation in the SARS-COV-2 Spike Protein.” medRxiv, January, 2021.01.12.20249080.

Harvey, William T., Alessandro M. Carabelli, Ben Jackson, Ravindra K. Gupta, Emma C. Thomson, Ewan M. Harrison, Catherine Ludden, et al. 2021. “SARS-CoV-2 Variants, Spike Mutations and Immune Escape.” Nature Reviews. Microbiology 19 (7): 409–24.

Hill, V., L. Du Plessis, T. P. Peacock, D. Aggarwal, R. Colquhoun, A. M. Carabelli, N. Ellaby, et al. 2022. “The Origins and Molecular Evolution of SARS-CoV-2 Lineage B.1.1.7 in the UK.” Virus Evolution 8 (2). https://doi.org/10.1093/ve/veac080.

Hodcroft, Emma B., Moira Zuber, Sarah Nadeau, Timothy G. Vaughan, Katharine H. D. Crawford, Christian L. Althaus, Martina L. Reichmuth, et al. 2021. “Spread of a SARS-CoV-2 Variant through Europe in the Summer of 2020.” Nature 595 (7869): 707–12.

Jones, Terry C., Guido Biele, Barbara Mühlemann, Talitha Veith, Julia Schneider, Jörn Beheim-Schwarzbach, Tobias Bleicker, et al. 2021. “Estimating Infectiousness throughout SARS-CoV-2 Infection Course.” Science, July. https://doi.org/10.1126/science.abi5273.

Katoh, Kazutaka, and Daron M. Standley. 2013. “MAFFT Multiple Sequence Alignment Software Version 7: Improvements in Performance and Usability.” Molecular Biology and Evolution 30 (4): 772–80.

Kidd, Michael, Alex Richter, Angus Best, Nicola Cumley, Jeremy Mirza, Benita Percival, Megan Mayhew, et al. 2021. “S-Variant SARS-CoV-2 Lineage B1.1.7 Is Associated With Significantly Higher Viral Load in Samples Tested by TaqPath Polymerase Chain Reaction.” The Journal of Infectious Diseases 223 (10): 1666–70.

Kraemer, Moritz U. G., D. A. T. Cummings, S. Funk, R. C. Reiner, N. R. Faria, O. G. Pybus, and S. Cauchemez. 2019. “Reconstruction and Prediction of Viral Disease Epidemics.” Epidemiology & Infection 147. https://doi.org/10.1017/S0950268818002881.

Kraemer, Moritz U. G., Verity Hill, Christopher Ruis, Simon Dellicour, Sumali Bajaj, John T. McCrone, Guy Baele, et al. 2021. “Spatiotemporal Invasion Dynamics of SARS-CoV-2 Lineage B.1.1.7 Emergence.” Science 373 (6557): 889–95.

Lemey, Philippe, Nick Ruktanonchai, Samuel L. Hong, Vittoria Colizza, Chiara Poletto, Frederik Van den Broeck, Mandev S. Gill, et al. 2021. “Untangling Introductions and Persistence in COVID-19 Resurgence in Europe.” Nature 595 (7869): 713–17.

Lythgoe, Katrina A., Matthew Hall, Luca Ferretti, Mariateresa de Cesare, George MacIntyre- Cockett, Amy Trebes, Monique Andersson, et al. 2021. “SARS-CoV-2 within-Host Diversity and Transmission.” Science 372 (6539). https://doi.org/10.1126/science.abg0821.

Markov, Peter V., Mahan Ghafari, Martin Beer, Katrina Lythgoe, Peter Simmonds, Nikolaos I. Stilianakis, and Aris Katzourakis. 2023. “The Evolution of SARS-CoV-2.” Nature Reviews. Microbiology, April, 1–19.

McCarthy, Kevin R., Linda J. Rennick, Sham Nambulli, Lindsey R. Robinson-McCarthy, William G. Bain, Ghady Haidar, and W. Paul Duprex. 2021. “Recurrent Deletions in the SARS-CoV-2 Spike Glycoprotein Drive Antibody Escape.” Science 371 (6534): 1139– 42.

Meng, Bo, Steven A. Kemp, Guido Papa, Rawlings Datir, Isabella A. T. Ferreira, Sara Marelli, William T. Harvey, et al. 2021. “Recurrent Emergence of SARS-CoV-2 Spike Deletion H69/V70 and Its Role in the Alpha Variant B.1.1.7.” Cell Reports 35 (13). https://doi.org/10.1016/j.celrep.2021.109292.

Minh, Bui Quang, Heiko A. Schmidt, Olga Chernomor, Dominik Schrempf, Michael D. Woodhams, Arndt von Haeseler, and Robert Lanfear. 2020. “IQ-TREE 2: New Models and Efficient Methods for Phylogenetic Inference in the Genomic Era.” Molecular Biology and Evolution 37 (5): 1530–34.

Mohanan, Manoj, Anup Malani, Kaushik Krishnan, and Anu Acharya. 2021. “Prevalence of SARS-CoV-2 in Karnataka, India.” JAMA: The Journal of the American Medical Association 325 (10): 1001–3.

Neher, Richard A. 2022. “Contributions of adaptation and purifying selection to SARS-CoV-2 Evolution”. Virus Evolution 8 (2). https://doi.org/10.1093/ve/veac113

Office for National Statistics. 2021a. “Coronavirus (COVID-19) Infection Survey QMI.” 2021. https://www.ons.gov.uk/peoplepopulationandcommunity/healthandsocialcare/conditionsanddiseases/methodologies/coronaviruscovid19infectionsurveyqmi.

Office for National Statistics. 2021b. “Coronavirus (COVID-19) Infection Survey: Technical Data - Office for National Statistics.” 2021. https://www.ons.gov.uk/peoplepopulationandcommunity/healthandsocialcare/conditionsanddiseases/datasets/covid19infectionsurveytechnicaldata/2021.

Office for National Statistics. 2021c. “Coronavirus (COVID-19) Infection Survey, UK - Office for National Statistics.” Office for National Statistics. 2021. https://www.ons.gov.uk/peoplepopulationandcommunity/healthandsocialcare/conditionsanddiseases/bulletins/coronaviruscovid19infectionsurveypilot/16july2021.

O’Toole, Áine, Emily Scher, Anthony Underwood, Ben Jackson, Verity Hill, John T. McCrone, Rachel Colquhoun, et al. 2021. “Assignment of Epidemiological Lineages in an Emerging Pandemic Using the Pangolin Tool.” Virus Evolution 7 (2). https://doi.org/10.1093/ve/veab064.

Oude Munnink, Bas B., Nathalie Worp, David F. Nieuwenhuijse, Reina S. Sikkema, Bart Haagmans, Ron A. M. Fouchier, and Marion Koopmans. 2021. “The next Phase of SARS-CoV-2 Surveillance: Real-Time Molecular Epidemiology.” Nature Medicine 27 (9): 1518–24.

Pouwels, Koen B., Thomas House, Emma Pritchard, Julie V. Robotham, Paul J. Birrell, Andrew Gelman, Karina-Doris Vihta, et al. 2021. “Community Prevalence of SARS-CoV-2 in England from April to November, 2020: Results from the ONS Coronavirus Infection Survey.” The Lancet. Public Health 6 (1): e30–38.

Public Health England. 2021. “Variants of Concern Technical Briefing 10.” 2021. https://assets.publishing.service.gov.uk/government/uploads/system/uploads/attachment_data/file/984274/Variants_of_Concern_VOC_Technical_Briefing_10_England.pdf.

Rambaut, Andrew, Edward C. Holmes, Áine O’Toole, Verity Hill, John T. McCrone, Christopher Ruis, Louis du Plessis, and Oliver G. Pybus. 2020. “A Dynamic Nomenclature Proposal for SARS-CoV-2 Lineages to Assist Genomic Epidemiology.” Nature Microbiology 5 (11): 1403–7.

Rambaut, Andrew, N. Loman, O. Pybus, W. Barclay, J. Barrett, A. Carabelli, T. Connor, et al. 2020. “Preliminary Genomic Characterisation of an Emergent SARS-CoV-2 Lineage in the UK Defined by a Novel Set of Spike Mutations.” December 18, 2020. https://virological.org/t/preliminary-genomic-characterisation-of-an-emergent-sars-cov-2-lineage-in-the-uk-defined-by-a-novel-set-of-spike-mutations/563.

Rasmussen, Carl Edward, and Christopher K. I. Williams. 2005. “Gaussian Processes for Machine Learning.” https://doi.org/10.7551/mitpress/3206.001.0001.

Sagulenko, Pavel, Vadim Puller, and Richard A. Neher. 2018. “TreeTime: Maximum-Likelihood Phylodynamic Analysis.” Virus Evolution 4 (1). https://doi.org/10.1093/ve/vex042.

Sah, Pratha, Meagan C. Fitzpatrick, Charlotte F. Zimmer, Elaheh Abdollahi, Lyndon Juden- Kelly, Seyed M. Moghadas, Burton H. Singer, and Alison P. Galvani. 2021. “Asymptomatic SARS-CoV-2 Infection: A Systematic Review and Meta-Analysis.” Proceedings of the National Academy of Sciences of the United States of America 118 (34). https://doi.org/10.1073/pnas.2109229118.

Sonabend, R., L. K. Whittkles, N. Imai, P. N. Perez-Guzman, E. S. Knock, T. Rawson, K. A. M. Gaythorpe, et al. 2021. “Non-Pharmaceutical Interventions, Vaccination, and the SARS-CoV-2 Delta Variant in England: A Mathematical Modelling Study.” Lancet. https://doi.org/10.1016/S0140-6736(21)02276-5.

Subissi, Lorenzo, Anne von Gottberg, Lipi Thukral, Nathalie Worp, Bas B. Oude Munnink, Surabhi Rathore, Laith J. Abu-Raddad, et al. 2022. “An Early Warning System for Emerging SARS-CoV-2 Variants.” Nature Medicine 28 (6): 1110–15.

Tay, John H., Ashleigh F. Porter, Wytamma Wirth, and Sebastian Duchene. 2022. “The Emergence of SARS-CoV-2 Variants of Concern Is Driven by Acceleration of the Substitution Rate.” Molecular Biology and Evolution 39 (2): msac013.

Tegally, Houriiyah, Monika Moir, Josie Everatt, Marta Giovanetti, Cathrine Scheepers, Eduan Wilkinson, Kathleen Subramoney, et al. 2022. “Emergence of SARS-CoV-2 Omicron Lineages BA.4 and BA.5 in South Africa.” Nature Medicine, June, 1–6.

The COVID-19 Genomics UK (COG-UK) consortium. 2020. “An Integrated National Scale SARS-CoV-2 Genomic Surveillance Network.” The Lancet Microbe. https://doi.org/10.1016/S2666-5247(20)30054-9.

Thomson, Emma C., Laura E. Rosen, James G. Shepherd, Roberto Spreafico, Ana da Silva Filipe, Jason A. Wojcechowskyj, Chris Davis, et al. 2021. “Circulating SARS-CoV-2 Spike N439K Variants Maintain Fitness While Evading Antibody-Mediated Immunity.” Cell 184 (5): 1171–87.e20.

Turakhia, Yatish, Bryan Thornlow, Angie S. Hinrichs, Nicola De Maio, Landen Gozashti, Robert Lanfear, David Haussler, and Russell Corbett-Detig. 2021. “Ultrafast Sample Placement on Existing tRees (UShER) Enables Real-Time Phylogenetics for the SARS-CoV-2 Pandemic.” Nature Genetics 53 (6): 809–16.

UKHSA. 2021. “Surge Testing for New Coronavirus (COVID-19) Variants.” 2021. https://www.gov.uk/guidance/surge-testing-for-new-coronavirus-covid-19-variants.

UKHSA. 2023. Genome Sequence Prevalence and Growth Rate Update: 24 May 2023.” GOV.UK. Accessed June 2, 2023. https://www.gov.uk/government/publications/sars-cov-2-genome-sequence-prevalence-and-growth-rate/sars-cov-2-genome-sequence-prevalence-and-growth-rate-update-24-may-2023.

Viana, Raquel, Sikhulile Moyo, Daniel G. Amoako, Houriiyah Tegally, Cathrine Scheepers, Christian L. Althaus, Ugochukwu J. Anyaneji, et al. 2021. “Rapid Epidemic Expansion of the SARS-CoV-2 Omicron Variant in Southern Africa.” medRxiv, December, 2021.12.19.21268028.

Vogels, Chantal B. F., Mallery I. Breban, Isabel M. Ott, Tara Alpert, Mary E. Petrone, Anne E. Watkins, Chaney C. Kalinich, et al. 2021. “Multiplex qPCR Discriminates Variants of Concern to Enhance Global Surveillance of SARS-CoV-2.” PLoS Biology 19 (5): e3001236.

Vöhringer, Harald S., Theo Sanderson, Matthew Sinnott, Nicola De Maio, Thuy Nguyen, Richard Goater, Frank Schwach, et al. 2021. “Genomic Reconstruction of the SARS-CoV-2 Epidemic in England.” Nature, October, 1–11.

Volz, Erik, Swapnil Mishra, Meera Chand, Jeffrey C. Barrett, Robert Johnson, Lily Geidelberg, Wes R. Hinsley, et al. 2021. “Assessing Transmissibility of SARS-CoV-2 Lineage B.1.1.7 in England.” Nature 593 (7858): 266–69.

Walker, Ann Sarah, Karina-Doris Vihta, Owen Gethings, Emma Pritchard, Joel Jones, Thomas House, Iain Bell, et al. 2021. “Increased Infections, but Not Viral Burden, with a New SARS-CoV-2 Variant.” medRxiv, January, 2021.01.13.21249721.

WHO. 2023. WHO Global Genomic Surveillance Strategy for Pathogens with Pandemic and Epidemic Potential 2022-2032.” n.d. Accessed June 2, 2023. https://www.who.int/initiatives/genomic-surveillance-strategy.

WHO. 2023. “Tracking SARS-CoV-2 Variants.” Accessed April 5, 2023. https://www.who.int/activities/tracking-SARS-CoV-2-variants.

Wu, Sean L., Andrew N. Mertens, Yoshika S. Crider, Anna Nguyen, Nolan N. Pokpongkiat, Stephanie Djajadi, Anmol Seth, et al. 2020. “Substantial Underestimation of SARS-CoV-2 Infection in the United States.” Nature Communications 11 (1): 1–10.

Wymant, Chris, François Blanquart, Tanya Golubchik, Astrid Gall, Margreet Bakker, Daniela Bezemer, Nicholas J. Croucher, et al. 2018. “Easy and Accurate Reconstruction of Whole HIV Genomes from Short-Read Sequence Data with Shiver.” Virus Evolution 4 (1): vey007.

Yu, Guangchuang, David K. Smith, and Zhu Huachen Lam Tsan-Yuk. 2017. “Ggtree: An R Package for Visualization and Annotation of Phylogenetic Trees with Their Covariates and Other Associated Data.” Methods in Ecology and Evolution 8: 28–36.

Zhao, Lei, and Christopher J. R. Illingworth. 2019. “Measurements of Intrahost Viral Diversity Require an Unbiased Diversity Metric.” Virus Evolution 5 (1): vey041.

